# Protection against Omicron conferred by mRNA primary vaccine series, boosters, and prior infection

**DOI:** 10.1101/2022.05.26.22275639

**Authors:** Elizabeth T. Chin, David Leidner, Lauren Lamson, Kimberley Lucas, David M. Studdert, Jeremy D. Goldhaber-Fiebert, Jason R. Andrews, Joshua A. Salomon

**Author notes:** **Corresponding Author:** Dr. Elizabeth T. Chin, Medical School Office Building, 1265 Welch Road, Stanford, CA 94305.

## Abstract

**Background:** Prisons and jails are high-risk settings for Covid-19 transmission, morbidity, and mortality. We evaluate protection conferred by prior infection and vaccination against the SARS-CoV-2 Omicron variant within the California state prison system.

**Methods:** We employed a test-negative design to match resident and staff cases during the Omicron wave (December 24, 2021—April 14, 2022) to controls according to a case’s test-week as well as demographic, clinical, and carceral characteristics. We estimated protection against infection using conditional logistic regression, with exposure status defined by vaccination, stratified by number of mRNA doses received, and prior infection, stratified by periods before or during Delta variant predominance.

**Results:** We matched 15,783 resident and 8,539 staff cases to 180,169 resident and 90,409 staff controls. Among cases, 29.7% and 2.2% were infected before or during the emergence of the Delta variant, respectively; 30.6% and 36.3% were vaccinated with two or three doses, respectively. Estimated protection from Omicron infection for two and three doses were 14.9% (95% Confidence Interval [CI], 12.3—19.7%) and 43.2% (42.2—47.4%) for those without known prior infections, 47.8% (95% CI, 46.6—52.8%) and 61.3% (95% CI, 60.7—64.8%) for those infected before the emergence of Delta, and 73.1% (95% CI, 69.8—80.1%) and 86.8% (95% CI, 82.1—92.7) for those infected during the period of Delta predominance.

**Conclusion:** A third mRNA dose provided significant, additional protection over two doses, including among individuals with prior infection. Our findings suggest that vaccination should remain a priority—even in settings with high levels of transmission and prior infection.

---

The BNT162b2 (Pfizer-BioNTech) and mRNA-1273 (Moderna) vaccines have been highly effective in protecting against severe acute respiratory syndrome coronavirus 2 (SARS-CoV-2) infection and coronavirus disease 2019 (Covid-19) illness. Evidence of effectiveness comes largely from studies of the ancestral strain and variants prior to the B.1.1.529 (Omicron) variant^1–4^. Prior infection also confers protection against reinfection^1,5^. However, both infection- and vaccine-acquired protection against infection wane over time^6–9^.

Recent studies have shown continued effectiveness of vaccination against hospitalizations or death^10–13^, though reduced effectiveness for confirmed^11,14^ and symptomatic illness^10,15,16^ with Omicron. While some studies have reported estimates of hybrid immunity against infection with Omicron^10,14^, information among individuals with recent and remote prior infections remains limited.

In this study, we analyzed data from California Department of Corrections and Rehabilitation (CDCR), which operates the second largest state prison system in the United States. Prisons and jails are especially risky congregate settings for Covid-19, and have been the sites of many large outbreaks during the pandemic^1,17^. CDCR began offering third mRNA vaccine doses to resident and staff at the end of August 2021, with 54.1% of residents and 20.2% of prison staff boosted by the end of December 2021. The Omicron variant was first identified within the CDCR system among assayed positive samples collected from correctional staff on December 10, 2021, with substantial outbreaks emerging among both residents and staff shortly after, consistent with the broader global wave of Omicron infection.

We analyzed confirmed SARS-CoV-2 infections among nearly 70,000 incarcerated people and over 20,000 correctional staff in California during the Omicron wave (December 24, 2021 through April 14, 2022) to estimate the effectiveness of mRNA vaccines against infection, stratified by number of doses received, and by prior infection, stratified by periods before or during predominance of the Delta variant.

## Methods

### Study design and sample

We conducted a retrospective test-negative design^18^ study starting December 24, 2021, two weeks after Omicron was initially identified, and spanning the 16-week period until April 14, 2022, during which the Omicron variant dominated SARS-CoV-2 infections detected in and around California state prisons^19^. Our study analyzed two high-risk populations: residents and correctional staff at these prisons.

Residents were eligible for inclusion in the present study if they were incarcerated in a CDCR prison during the study period. To ensure a focus on staff at highest risk of workplace exposures, we included staff members who worked in custody or healthcare positions (excluding contract employees), had regular direct contact with residents in those job roles, and worked at least half of the number of days in the study period (≥56 shifts).

Next, residents and staff meeting the above criteria were excluded if they were not tested during the study period, received an Ad26.COV2.S (Janssen) vaccine, were vaccinated with one mRNA vaccine dose, received a vaccine not approved or authorized by the U.S. Food and Drug Administration, received both BNT162b2 (Pfizer-BioNTech) and mRNA-1273 (Moderna) vaccines, had off-schedule vaccination^20^, or had incomplete demographic or locational data (Figure 1). To avoid misclassification of reinfections, we excluded tests for people who had a new documented infection within 90 days or any positive tests within 30 days prior to the test collection date. Given the latency of biologically plausible protection, we also excluded tests from people who had received a second or third vaccine dose within seven days leading up to the test collection date. We censored residents and staff after the date of test collection for a positive test or on the date of vaccination with a fourth mRNA dose.

**Figure 1.**
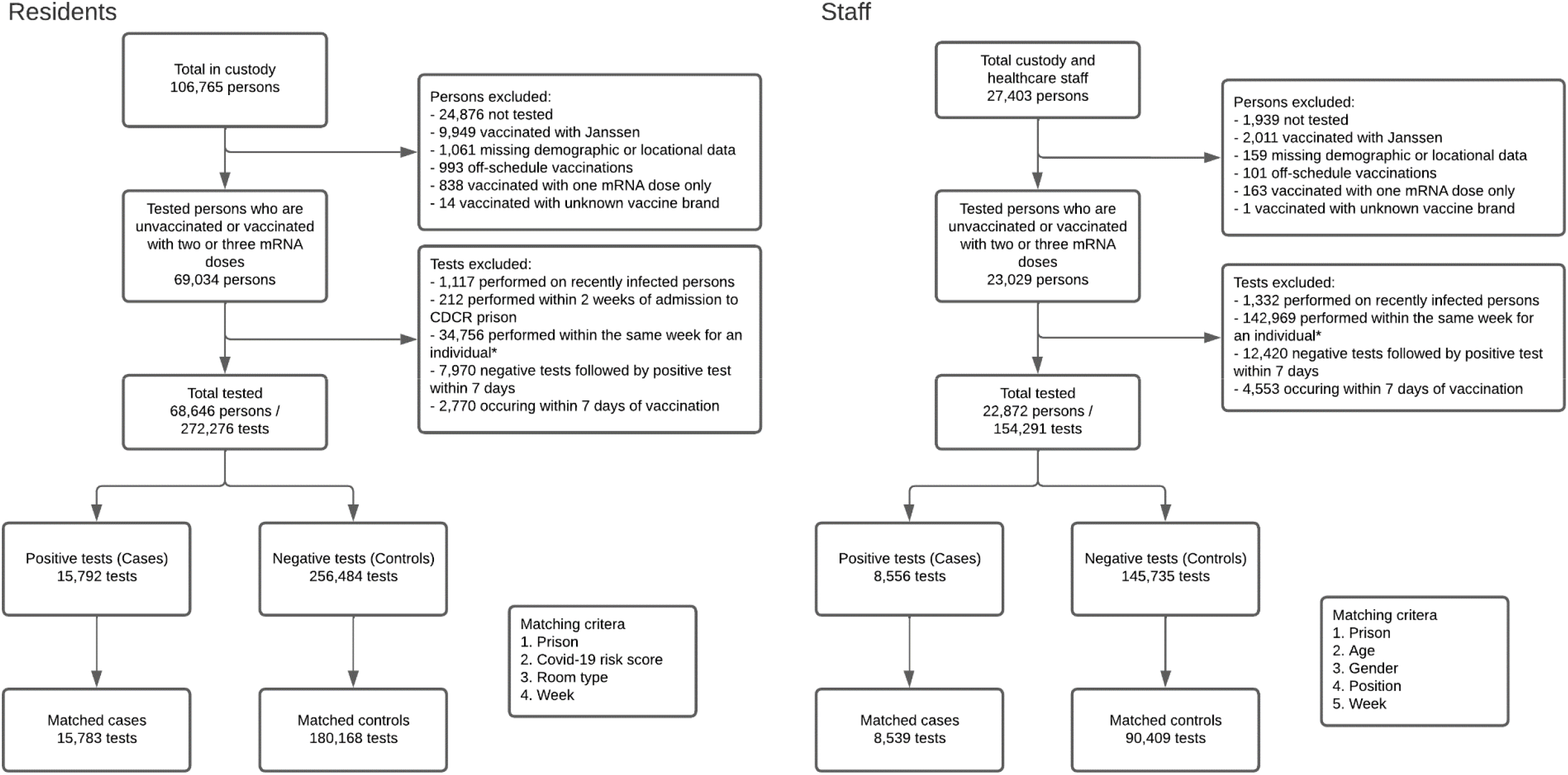
Flowchart of the study sample for both resident and staff populations, and the selection of cases and controls. ^**\***^ Controls were matched with cases by week. An individual could contribute a maximum of one test per week.

### Data and key measures

CDCR collects and stores daily de-identified data on all residents and staff. Detailed RT-PCR and antigen SARS-CoV-2 testing information for residents came from a multilayered voluntary testing program (99.9% RT-PCR) that included risk-based routine testing, surveillance testing, and testing in response to detected outbreaks (Table S1). All staff were tested through a twice-weekly mandatory RT-PCR testing program during outbreaks, defined as three or more related positive resident cases at a prison within a 14-day window. When prisons exited the outbreak phase, only staff who received fewer than two doses or were eligible for but not vaccinated with a third dose continued to undergo twice-weekly testing^21^. Voluntary testing was available for all staff, and close contacts of active cases were compulsorily tested^22^. By January 1, 2022, more than half of prisons had entered outbreak phase, with the last prison commencing on January 7, 2022.

We defined a prior SARS-CoV-2 infection as having at least one positive test in CDCR’s records prior to the beginning of the study period (Table S3 & Figure S3). Prior infections were stratified by those that occurred before and since July 1, 2021, reflecting periods of low and high B.1.617.2 (Delta) variant prevalence, respectively^1,19^. Among individuals with multiple prior infections, a person’s most recent prior infection determined their assigned category.

In addition to detailed information on testing and vaccination (e.g., dates, brand), the study data included sample members’ demographic characteristics (sex or gender, age, racial or ethnic group), residents’ carceral characteristics (prison, room type, security level), and staff’s carceral characteristics (prison, position type). For residents, documented history of 25 comorbid conditions (e.g., hypertension, chronic kidney disease, asthma) and a composite score designed by CDCR to grade risks of severe illness from SARS-CoV-2 infections and be used to guide Covid-19 mitigation policies (Table S2) were provided.

### Selection of cases and controls

Cases were defined as a resident or staff member who had a positive SARS-CoV-2 RT-PCR or antigen test. Controls had a negative SARS-CoV-2 test. We used coarsened exact matching^23^ to identify controls, matching to the case’s test week as well as several demographic, clinical, and carceral characteristics. Specifically, residents were matched on prison (from among 35 prisons), Covid-19 risk score (0, 1, ≥2), and room type (cell, dorm). Staff were matched on prison, position (custody, healthcare), age group (18-39, 40-54, ≥55), and gender (male, female). Residents were not matched on sex because men and women are generally housed in separate prisons, making this variable highly collinear with prison.

To avoid misclassification of cases as controls, we excluded negative tests occurring within 7 days prior to a positive test. Because the calendar week of testing was used as a matching criterion, sample members contributed no more than one observation per week.

### Statistical analysis

The main goal of our analysis was to estimate the protection against infection conferred by vaccination and by prior infection status. The primary outcome of interest was confirmed infection, and the primary exposures of interest were vaccination status (unvaccinated, two doses, three doses) and prior infection status (no known; infected prior to July 1, 2021; infected since July 1, 2021). We estimated vaccine- and infection-acquired protection (1-odds ratio) using weighted conditional logistic regression to account for the matched design, with a set of indicator variables describing combinations of vaccination and prior infection status. 95% confidence intervals were calculated using 10,000 bias-corrected accelerated bootstrapped samples for analyses using coarsened exact matching. We report results from analyses stratified by population (residents, staff), as well as in the two populations combined.

### Sensitivity analyses

We conducted four sets of sensitivity analyses. First, we estimated protection using a variation on the main model adjusting for mRNA vaccine product (BNT162b2, mRNA-1273). Second, we examined the sensitivity of estimates to our definition of recent infections, shortening the exclusion of prior infections from 90 to 30 days. Third, recognizing that our primary analysis mixes booster ineligible and booster eligible persons, we excluded tests from those vaccinated with two doses but not yet eligible for a third. Finally, to assess the sensitivity of our estimates to the matching method, we used propensity score matching^24^ as an alternative to coarsened exact matching for the pooled and stratified analyses.

All analyses were performed using R software, version 3.5.2 (R Foundation for Statistical Computing). Additional details regarding model and variable specifications are provided in the Supplementary Appendix.

### Study oversight

The study was approved by the institutional review board at Stanford University (protocol #55835). Results are reported in accordance with Strengthening the Reporting of Observational Studies in Epidemiology (STROBE) guidelines (checklist in Supplementary Appendix).^25^

## Results

### Sample characteristics

Among 68,646 residents and 22,872 staff who met the cohort inclusion criteria, 23.0% of residents and 37.4% of staff tested positive during the study period (Figure 1). The study sample consisted of 15,783 resident (99.9% of positive tests) and 8,539 staff (99.8% of positive tests) cases, matched with 180,169 resident (70.2% of negative tests) and 90,409 staff (62.0% of negative tests) controls. Staff had a higher weekly testing frequency over the study period, at 1.6 (sd: 0.5) compared to 0.8 (sd: 0.4) for residents (Table S3, Figures S2 & S3).

A third of cases had prior confirmed infections, with most occurring before the emergence of the Delta variant (Table S3). Among those infected prior to the emergence of the Delta variant, residents had a median of 421 days since a prior infection (Interquartile Range [IQR]: 395-478) compared to 406 days (IQR: 377-450) for staff.

Conversely, among those infected since the emergence of the Delta variant, residents had more recent infections, with a median days 150 days since a prior infection (IQR: 118-175) compared to 154 days (IQR: 127-181) for staff.

Coverage with at least the primary vaccine series was higher among residents (73.5% of cases) than staff (54.7% of staff). The difference in coverage was especially pronounced in relation to the third dose, with 49.1% of resident compared to only 12.7% of staff cases vaccinated with a booster. The staff sample had more recent vaccination across all vaccination statuses. Among the subset with two doses, the median days since last vaccination was 201 days (IQR: 109.3-331) for staff and 230 days (IQR: 111-310) for residents. Similarly, for the subset with three doses, the days since last vaccination was 59 days (IQR: 33-89) for staff and 68 days (IQR: 49-92) for residents.

### Protection due to vaccine- and infection-induced immunity

In the combined sample of residents and staff, vaccine effectiveness against confirmed SARS-CoV-2 infection for those without prior known infections was 14.9% (95% Confidence Interval [CI], 12.3—19.7%) for those vaccinated with two doses. Receiving a third dose had an estimated effectiveness of 33.2% (95% CI, 31.2—37.1%) compared to those vaccinated with two doses only, and 43.2% (42.2—47.4%) compared to the unvaccinated group (Table S4). Infection-conferred protection among the unvaccinated was 25.7% (95% CI, 23.4—31.6%) for those infected before the emergence of the Delta variant. Having a more recent infection, one after the emergence of Delta, had an estimated protection of 47.2% (95% CI, 42.2—54.3%) compared to infections before July 1, 2021 and 60.8% (58.4—66.8%) compared to no prior infections.

Across all prior infection statuses, a third dose provided significant, additional protection over two doses. For those infected before the emergence of the Delta variant, estimated protection was 47.8% (95% CI, 46.6—52.8%) and 61.3% (95% CI, 60.7—64.8%) for two and three doses, respectively. For those infected during the period of high Delta variant prevalence, estimated protection was 73.1% (95% CI, 69.8—80.1%) and 86.8% (95% CI, 82.1—92.7%) for two and three doses, respectively.

Relative levels of protection for vaccine and prior infection statuses for residents and staff were generally consistent (Figure 4, details in Table S4A and S5A).

**Figure 2.**
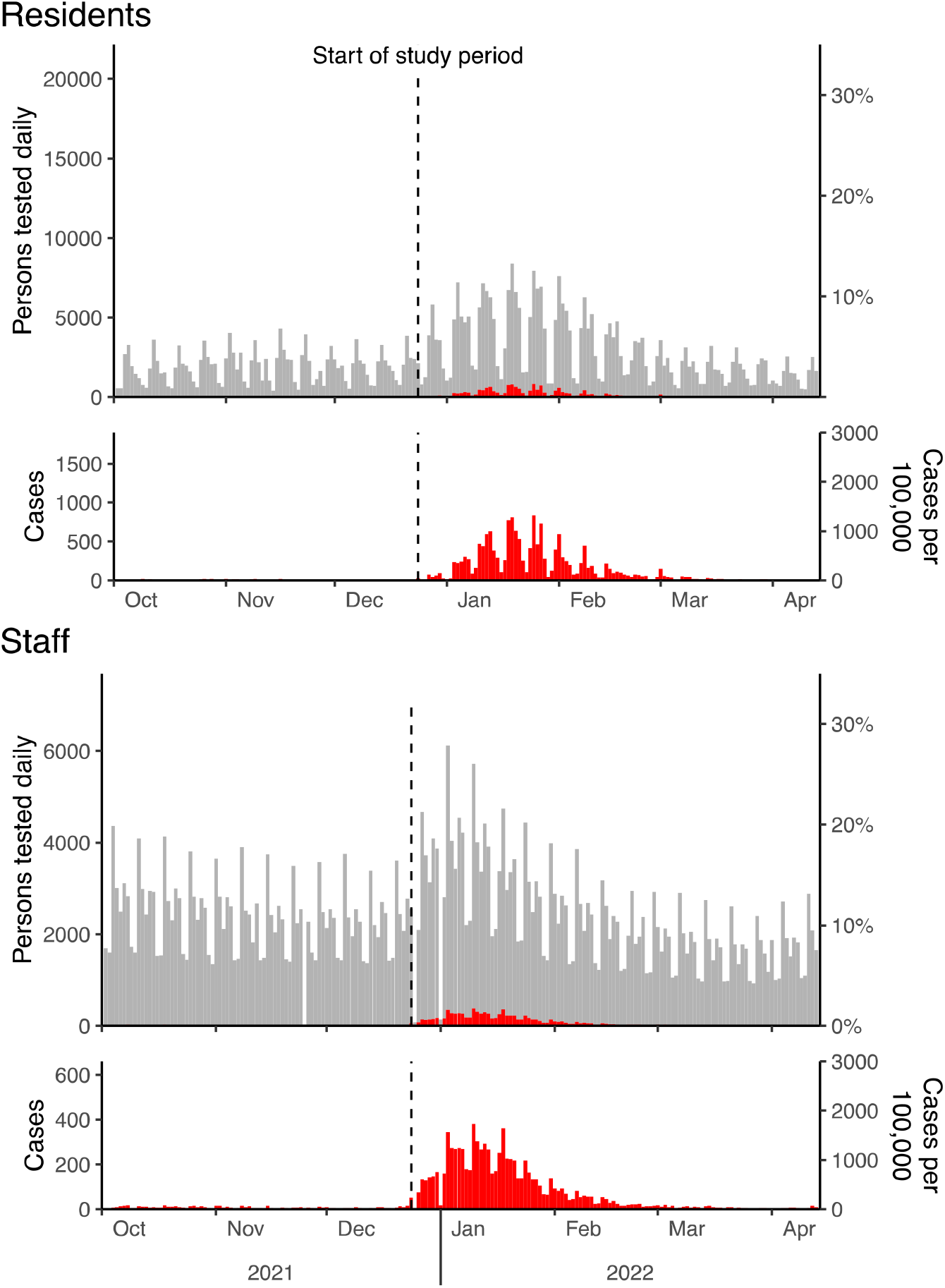
Testing and cases among study cohort. In the top panel, the daily number and percentage of residents and staff* tested and who were positive. In the bottom panel, the daily number of residents who tested positive. Testing and case series were extended over the historical period 2.5 months before the start of the study period. * Reduced staff testing on Federal holidays.

**Figure 3.**
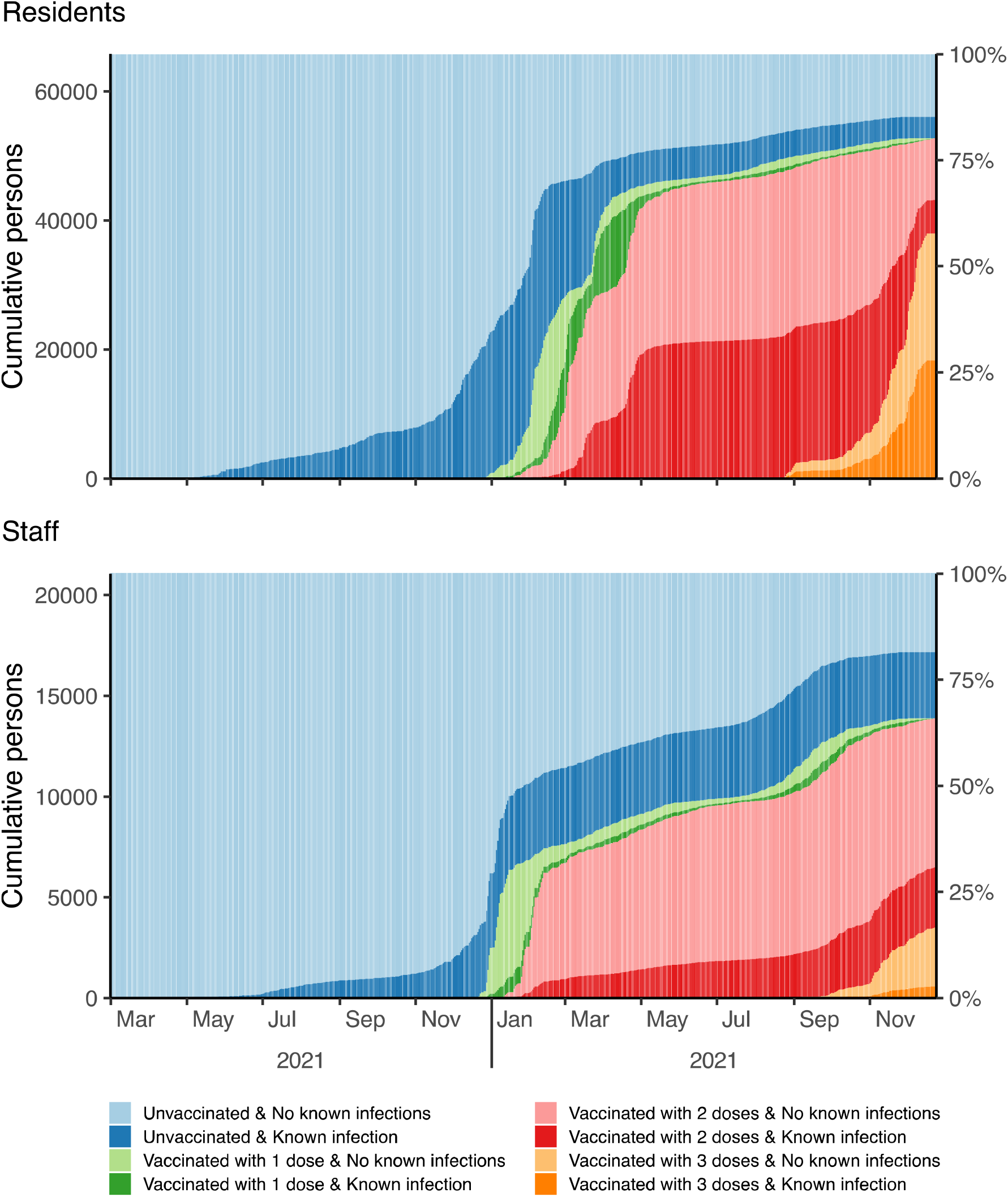
Vaccine and prior infection status of study cohort over time. Study cohort consisted of residents and staff who were unvaccinated or received mRNA vaccines for Covid-19.

**Figure 4.**
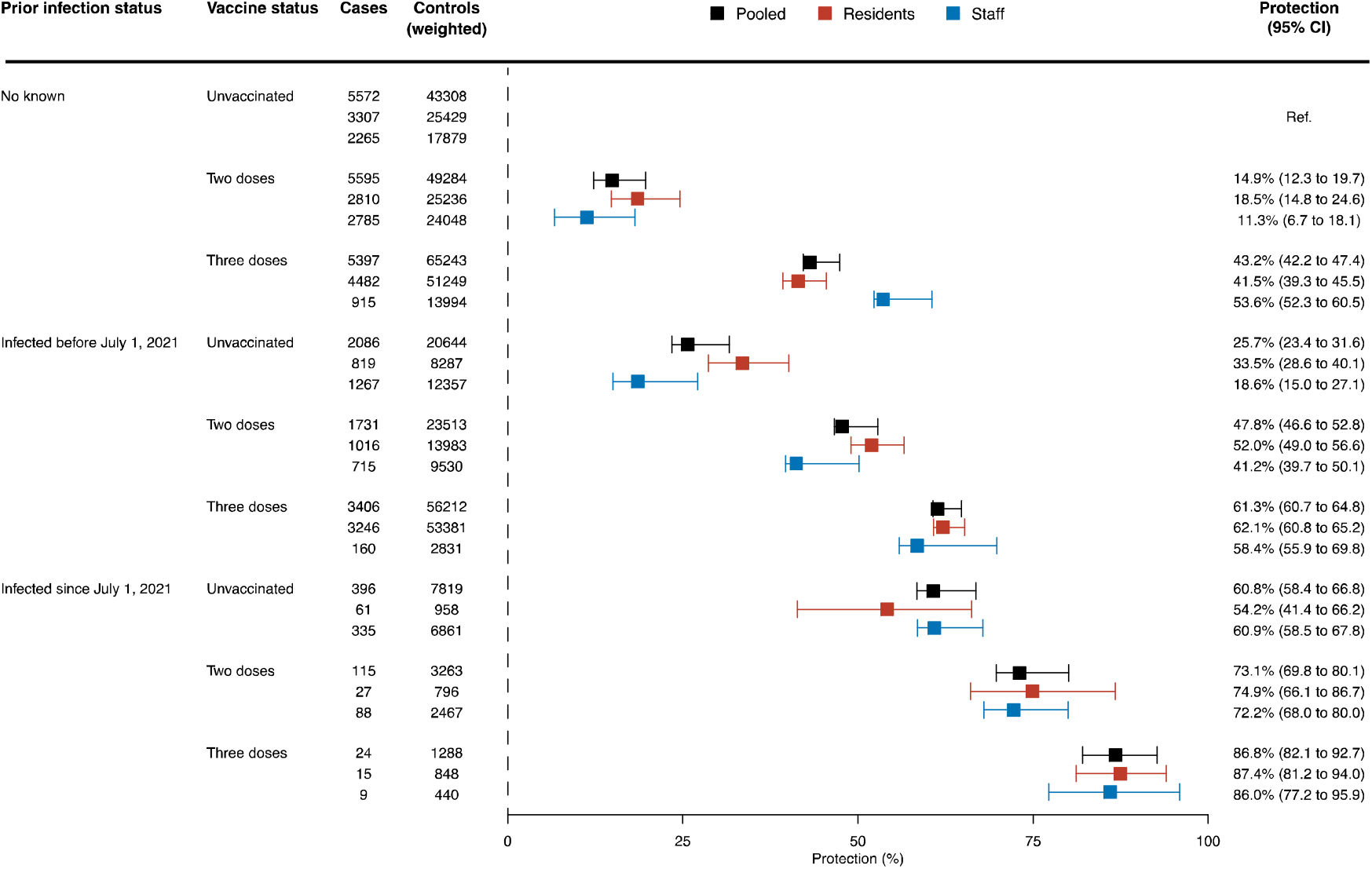
SARS-CoV-2 infections and adjusted estimates of effectiveness among incarcerated people and correctional staff in California prisons during Omicron Covid-19 outbreaks, December 24, 2021 to April 14, 2022. Individuals with vaccine statuses of two or three doses were vaccinated with mRNA vaccines. * Counts for controls were rescaled using weights derived from coarsened exact matching and sum to the total unweighted count. Weighted counts were rounded in this figure for display purposes.

### Sensitivity analyses

An alternative model specification adjusting for vaccine type produced similar estimates to the main analysis (Table S4A and S5A). Analyses in an expanded sample to include more recent prior infections resulted in slightly higher levels of protection for groups with infections since July 1, 2021 (Table S4B and S5B). In the analysis that excluded tests from persons not eligible for a third dose, estimates of protection for those vaccinated with two doses and who also did not have a recent prior infection were substantially lower than those in the main analysis (Table S4C and S5C). Results were consistent when using propensity score matching instead of coarsened exact matching, both for the main analysis and stratified analyses (Table S4D and S5D).

## Discussion

This study evaluated the protection conferred by two or three doses of BNT162b2 and mRNA-1273 vaccines and by prior infection during pre-Delta and Delta periods against confirmed SARS-CoV-2 infection from the Omicron variant among two high-risk populations. This study adds to the evidence base on vaccine effectiveness and immunity from prior infection by presenting a detailed comparison of infection risks across two categories defined in terms of both vaccination and infection status in populations with high levels of testing and presumed high ascertainment of infection status.

Compared to prior studies,^8,26–28^ our estimates for protection against reinfection were substantially lower than those analyzing infections during pre-Omicron outbreaks; furthermore, we found significant differences in protection conveyed by prior infections before and during the emergence of the Delta variant. Consistent with other studies^11,14^, we found the highest levels of protection among those who had three doses of vaccination and prior infections. Among those with prior infection, the incremental protection afforded by vaccination was substantial, especially vaccination with three doses, which raised protection against Omicron infection to levels similar to those reported in pre-Omicron studies of vaccination with the primary series.

Our study is one of few to report on the protection against Omicron conferred by three mRNA doses. Within groups without prior known infections, our estimates were similar to those in a convenience study of the effectiveness of two and three mRNA-1273 doses in a healthcare system in the United States^11^. Among people with prior infection, evidence of the additional benefit of a third dose is limited. A cohort study within another healthcare system in the United States did not find significant additional protection among those with prior infections for people vaccinated with three doses compared to people eligible for a third-dose^14^. Our main analysis of the primary vaccine series mixes both third-dose ineligible and third-dose eligible people, thus providing a potentially inflated estimate of effectiveness for those vaccinated with two doses only. Even with this conservative estimate, we observe significant, additional protection against confirmed infection from a third mRNA dose compared to two doses across both high-risk populations and all prior infection statuses.

To our knowledge, this is the first published study to assess both infection- and vaccine-acquired immunity for the Omicron variant with consideration to the timing of prior infections. It has several strengths. We used detailed daily information on vaccination status and key Covid-19 outcomes for each member of these two high-risk populations. These data allowed us to adjust for key potential confounders, demographic and exposure-related characteristics. An extensive testing program in both populations facilitated relatively complete measurement of SARS-CoV-2 infections. Large sample sizes in two distinct populations enabled relatively high precision in our estimates, and the similarity of estimates across the two populations is notable despite their different living situations, testing programs, and population structure.

Our study also has several limitations. As an observational cohort study, potential for bias due to confounding is an important consideration. While we aimed to limit confounding by matching on a variety of covariates, including those related to vaccine acceptance and risk of prior infections, the potential for confounding from unmeasured covariates remains. Vaccine uptake varied between residents and staff, and there were differences in the timing of uptake between populations. Moreover, there were differences across the two populations in the timing of prior infections. Differences between the two populations in relative infection risks by vaccination and prior infection status may in part reflect complex interactions of vaccine and prior infection timing, as could the order of vaccination and prior infection among those with both vaccine and infection-acquired immunity. Furthermore, CDCR conducted limited viral sequencing or molecular testing historically and during the study period, and thus we cannot disentangle the effects of variants from temporal waning, nor confirm that all cases observed during the study period were Omicron infections.

One potential violation of the assumptions in the test-negative design that testing was not compulsory for all staff during the entire study period. Estimates derived from staff who were boosted or vaccinated with two doses but ineligible for boosters may be biased downward since those staff were no longer required to undergo routine testing as cases declined. Additionally, testing for residents was neither routine, random, nor compulsory, potentially violating the test-negative design assumptions if testing motivation was mediated by vaccination and prior infection status. Several results provide reassurance. First, while staff testing was uneven across vaccination and prior infection statuses, staff were tested at least once weekly (range of means: 1.2—1.8 tests per week), providing relatively complete case detection. Furthermore, this bias is expected to be limited since nearly 90% of cases occurred during the outbreak phase when routine testing was mandatory for all staff. Second, while residents were less frequently tested than staff, testing was relatively even across vaccination and prior infection statuses (range of means: 0.7—0.8 tests per week). Most importantly, the consistency of the stratified analyses in these two populations with different testing procedures and exposures provide some confidence that bias related to variation in testing practices may not be of major concern.

Two additional limitations are noted. First, our estimates of protection focused on confirmed infections, not other important outcomes, such as symptomatic infections or severe disease. Incidence of hospitalizations and deaths in our sample was too low to support rigorous analysis of those outcomes within the vaccine and prior infection statuses examined, and symptom reporting was unreliable during the study period.^29^ Finally, the generalizability of our results to jails and other correctional systems, other high-risk populations (e.g., residents of skilled nursing facilities, healthcare workers, residents of homeless shelters), and lower-risk populations is unknown.

Findings from this study suggest that while mRNA vaccines and prior infections provide less protection from the Omicron variant compared to prior variants, boosters continue to provide significant additional protection, including among recently infected people. Continued emphasis on vaccination and other ongoing mitigation practices are essential in preventing transmission in populations that have already disproportionately borne a large share of disease burden during the Covid-19 pandemic.

## Data Availability

Data not available due to Data Use Agreement between Stanford University and the California Department of Corrections and Rehabilitation.

## ACKNOWLEDGMENTS

We thank Joseph Bick, John Dunlap, Heidi Bauer and the other staff members at California Department of Corrections and Rehabilitation for providing data and assistance with interpretation of study results. We also acknowledge help from members of the Stanford-CIDE Coronavirus Simulation Model (SC-COSMO) consortium.

## AUTHORS’ CONTRIBUTIONS

JAS, ETC, JRA, DMS, JDGF contributed to study design. ETC, DL, LL contributed to data analysis. All authors contributed to data interpretation. ETC, JAS wrote the first draft of the manuscript. JAS, ETC, JRA, DMS, JDGF, DL, LL, KL reviewed and edited the manuscript. All authors approved the final draft.

## NOTES

Potential conflicts of interest. All authors: No reported conflicts of interest. All authors have submitted the ICMJE Form for Disclosure of Potential Conflicts of Interest. Conflicts that the editors consider relevant to the content of the manuscript have been disclosed.

---

**Table 1.**
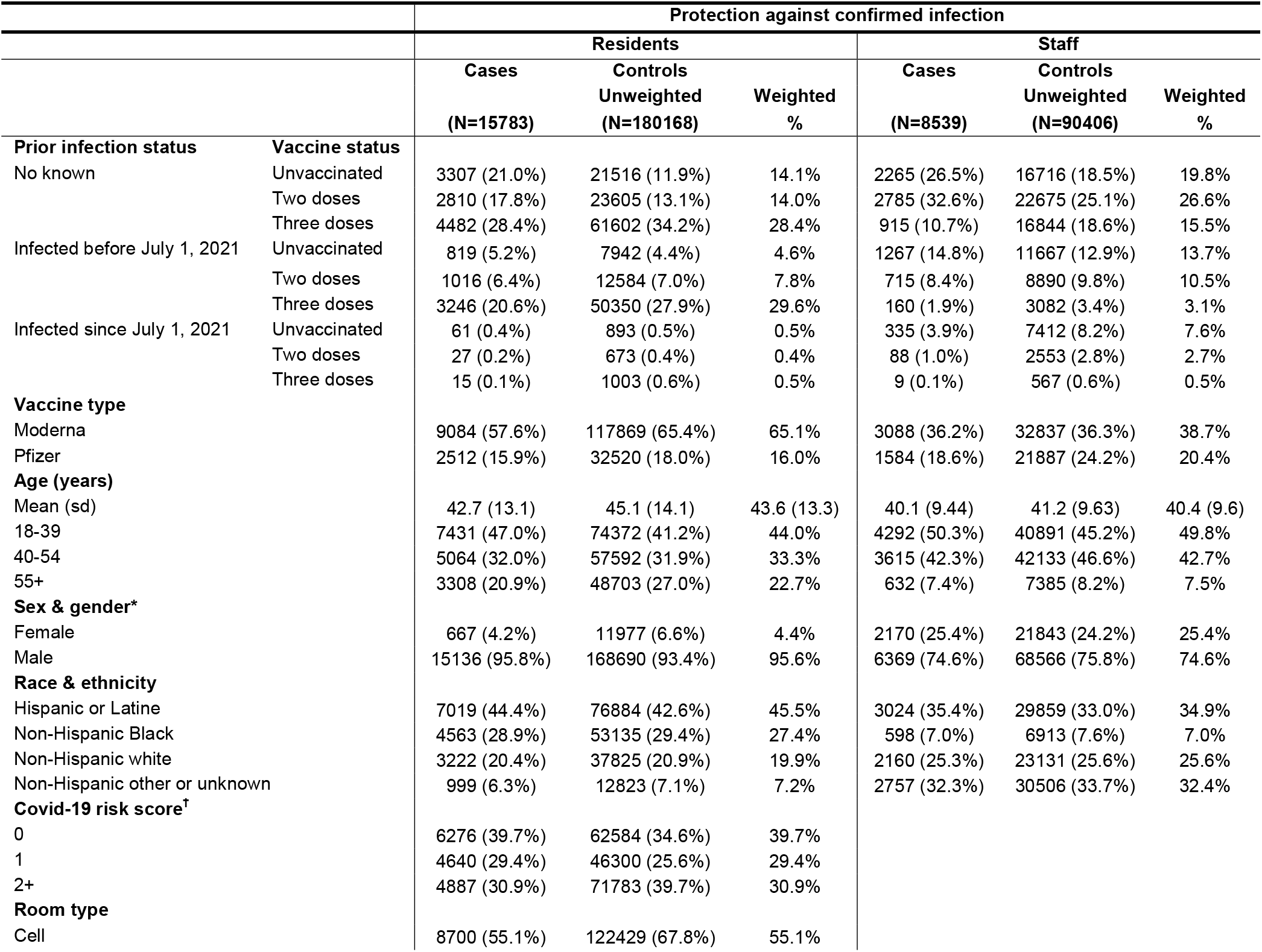
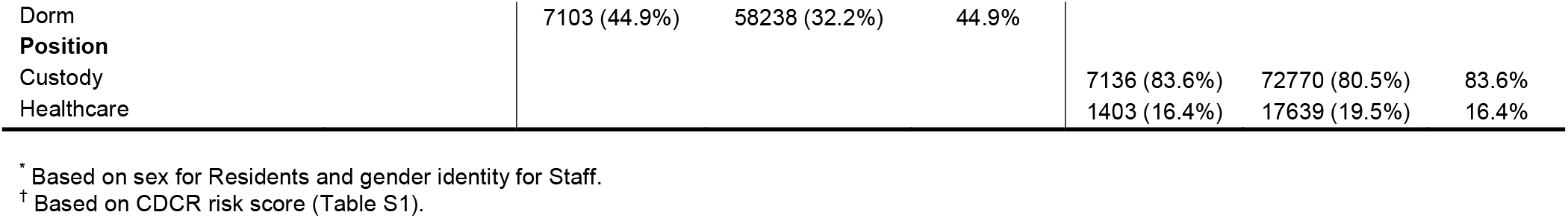
Characteristics of cases and controls. Demographic, health, and carceral characteristics of the matched study sample.

## REFERENCES

1. Chin ET, Leidner D, Zhang Y, et al. Effectiveness of the mRNA-1273 Vaccine during a SARS-CoV-2 Delta Outbreak in a Prison. N Engl J Med 2021;385(24):2300–1.

2. Chin ET, Leidner D, Zhang Y, et al. Effectiveness of COVID-19 vaccines among incarcerated people in California state prisons: retrospective cohort study. Clin Infect Dis 2022;ciab1032.

3. Dagan N, Barda N, Kepten E, et al. BNT162b2 mRNA Covid-19 Vaccine in a Nationwide Mass Vaccination Setting. N Engl J Med 2021;384(15):1412–23.

4. Lopez Bernal J, Andrews N, Gower C, et al. Effectiveness of Covid-19 Vaccines against the B.1.617.2 (Delta) Variant. New England Journal of Medicine 2021;385(7):585–94.

5. Baden LR, El Sahly HM, Essink B, et al. Efficacy and Safety of the mRNA-1273 SARS-CoV-2 Vaccine. New England Journal of Medicine 2021;384(5):403–16.

6. Abu-Raddad LJ, Chemaitelly H, Bertollini R. Waning mRNA-1273 Vaccine Effectiveness against SARS-CoV-2 Infection in Qatar. New England Journal of Medicine 2022;386(11):1091–3.

7. Nordström P, Ballin M, Nordström A. Risk of infection, hospitalisation, and death up to 9 months after a second dose of COVID-19 vaccine: a retrospective, total population cohort study in Sweden. The Lancet 2022;399(10327):814–23.

8. Hall V, Foulkes S, Insalata F, et al. Protection against SARS-CoV-2 after Covid-19 Vaccination and Previous Infection. N Engl J Med 2022;386(13):1207–20.

9. Chemaitelly H, Tang P, Hasan MR, et al. Waning of BNT162b2 Vaccine Protection against SARS-CoV-2 Infection in Qatar. New England Journal of Medicine 2021;385(24):e83.

10. Altarawneh HN, Chemaitelly H, Ayoub H, et al. Effect of prior infection, vaccination, and hybrid immunity against symptomatic BA.1 and BA.2 Omicron infections and severe COVID-19 in Qatar. Preprint. May 1 2022. https://www.medrxiv.org/content/10.1101/2022.03.22.22272745v1.

11. Tseng HF, Ackerson BK, Luo Y, et al. Effectiveness of mRNA-1273 against SARS-CoV-2 Omicron and Delta variants. Nat Med 2022;1–9.

12. Thompson MG, Natarajan K, Irving SA, et al. Effectiveness of a Third Dose of mRNA Vaccines Against COVID-19-Associated Emergency Department and Urgent Care Encounters and Hospitalizations Among Adults During Periods of Delta and Omicron variant Predominance - VISION Network, 10 States, August 2021-January 2022. MMWR Morb Mortal Wkly Rep 2022;71(4):139–45.

13. Johnson AG, Amin AB, Ali AR, et al. COVID-19 Incidence and Death Rates Among Unvaccinated and Fully Vaccinated Adults with and Without Booster Doses During Periods of Delta and Omicron variant Emergence - 25 U.S. Jurisdictions, April 4-December 25, 2021. MMWR Morb Mortal Wkly Rep 2022;71(4):132–8.

14. Lind ML, Robertson AJ, Silva J, et al. Effectiveness of Primary and Booster COVID-19 mRNA Vaccination against Infection Caused by the SARS-CoV-2 Omicron variant in People with a Prior SARS-CoV-2 Infection. Preprint. May 1 2022. https://www.medrxiv.org/content/10.1101/2022.04.19.22274056v2.

15. Andrews N, Stowe J, Kirsebom F, et al. Covid-19 Vaccine Effectiveness against the Omicron (B.1.1.529) Variant. N Engl J Med 2022;386(16):1532–46.

16. Accorsi EK, Britton A, Fleming-Dutra KE, et al. Association Between 3 Doses of mRNA COVID-19 Vaccine and Symptomatic Infection Caused by the SARS-CoV-2 Omicron and Delta variants. JAMA 2022;327(7):639–51.

17. Chin ET, Ryckman T, Prince L, et al. COVID-19 in the California State Prison System: an Observational Study of Decarceration, Ongoing Risks, and Risk Factors. J Gen Intern Med 2021;36(10):3096–102.

18. Jackson ML, Nelson JC. The test-negative design for estimating influenza vaccine effectiveness. Vaccine 2013;31(17):2165–8.

19. California’s COVID-19 response. Variants in California. Website. May 1, 2022. https://covid19.ca.gov/variants/.

20. Centers for Disease Control and Prevention. Interim COVID-19 Immunization Schedule for Ages 5 Years and Older. Updated April 23 2022. CS321629-AV. https://www.cdc.gov/vaccines/covid-19/downloads/COVID-19-immunization-schedule-ages-5yrs-older.pdf.

21. California Department of Corrections and Rehabiliation. Mandatory COVIDLJ19 Vaccines and Testing for Institution Staff. Updated August 19 2021. https://www.cdcr.ca.gov/covid19/mandatory-covid-19-vaccination-booster-and-testing-for-institution-facility-staff/.

22. California Department of Corrections and Rehabilitation. COVIDLJ19 Response Efforts. Updated February 2 2022. https://www.cdcr.ca.gov/covid19/covid-19-response-efforts/.

23. Iacus SM, King G, Porro G. Causal Inference without Balance Checking: Coarsened Exact Matching. Political Analysis 2012;20(1):1–24.

24. Ho DE, Imai K, King G, Stuart EA. Matching as Nonparametric Preprocessing for Reducing Model Dependence in Parametric Causal Inference. Political Analysis 2007;15(3):199–236.

25. von Elm E, Altman DG, Egger M, et al. The Strengthening the Reporting of Observational Studies in Epidemiology (STROBE) statement: guidelines for reporting observational studies. Lancet 2007;370(9596):1453–7.

26. Gazit S, Shlezinger R, Perez G, et al. SARS-CoV-2 Naturally Acquired Immunity vs. Vaccine-induced Immunity, Reinfections versus Breakthrough Infections: a Retrospective Cohort Study. Clin Infect Dis 2022;ciac262.

27. Shrestha NK, Burke PC, Nowacki AS, Terpeluk P, Gordon SM. Necessity of Coronavirus Disease 2019 (COVID-19) Vaccination in Persons Who Have Already Had COVID-19. Clinical Infectious Diseases 2022;ciac022.

28. Sheehan MM, Reddy AJ, Rothberg MB. Reinfection Rates Among Patients Who Previously Tested Positive for Coronavirus Disease 2019: A Retrospective Cohort Study. Clin Infect Dis 2021;73(10):1882–6.

29. Ryckman T, Chin ET, Prince L, et al. Outbreaks of COVID-19 variants in US prisons: a mathematical modelling analysis of vaccination and reopening policies. Lancet Public Health 2021;6(10):e760–70.

