## Supplemental Appendix for "Protection against Omicron conferred by mRNA primary vaccine series, boosters, and prior infection"

**SUPPLEMENTARY APPENDIX**

### 1 Investigators

***Stanford University***

Elizabeth T. Chin

Lauren Lamson

David M. Studdert

Jeremy D. Goldhaber-Fiebert

Jason R. Andrews

Joshua A. Salomon

***California Department of Corrections and Rehabilitation***

David Leidner

***California Correctional Health Care Services***

Kimberley Lucas

### 2 Study design

#### 2.1 Study setting

The California Department of Corrections and Rehabilitation (CDCR) provided de-identified person-day level data. The data included comprehensive information on PCR and antigen testing and vaccination for both prison staff and incarcerated residents (March 21, 2020 through April 14, 2022) across CDCR’s 35 prisons.^[1]^

During the outbreak, prisons implemented infection control measures including but not limited to restrictions on resident movement, quarantine, isolation, Covid-19 response testing, cleaning of areas of potential transmission, suspension of in-person and family visiting, staffing of essential personnel only, random audits of masking compliance, closing outdoor telephone booths, and programming with reduced group sizes, modified hours, staggered schedules, or in non-traditional spaces to allow for physical distancing.

#### 2.2 Definition of the analytic sample

Daily data extracts provided by the California Department of Corrections and Rehabilitation (CDCR) included a unique pseudo-identifier that allowed us to follow residents and staff over time.

Residents were included in the analytic sample if they:

- were incarcerated in a CDCR prison between December 24, 2021 and April 14, 2022

Staff were included in the analytic sample if they:

- were employed as custody or healthcare staff
- were employed in roles involving direct contact with residents
- worked at least 56 shifts during the study period

We excluded:

- tests before December 24, 2021 or after April 14, 2022
- individuals who were not tested
- individuals vaccinated with the Ad26.COV2.S vaccine
- individuals with partial vaccination status, defined as the day an individual receives their first vaccine dose to 7 days after they receive a second dose or days 0 to 7 after they receive a third dose
- individuals with off-schedule vaccinations as per recommended protocol^[2]^
- individuals with incomplete prison, Covid-19 risk score, or security level data for residents and prison and gender data for staff
- negative tests followed by a positive test within 7 days (to avoid misclassification of cases as controls)
- tests occurring on or after date of vaccination
- tests occurring after a positive test
- multiple tests collected from the same person on the same week, such that a maximum of one test per week was included per person.

#### 2.3 Study period

The study began two weeks after the first omicron case was detected in the CDCR system. The study period consisted of the 16-week period from December 24, 2021 to April 14, 2022.

#### 2.4 Variables

*Prior SARS-CoV-2 infection*. The data included Covid-19 testing information. CDCR has undertaken extensive testing of residents and staff for SARS-CoV-2 since April 2020, using real-time PCR and antigen tests (Figure S1). We defined a prior SARS-CoV-2 infection as having had at least one positive test in CDCR’s clinical records prior to the beginning of the study period (Table S3 & Figure S3). Dates were calculated as the date of test collection for the start of detection of a prior infection. Only the most recent prior infection was included. Prior infections were split into two categories, infections that occurred prior to July 1, 2021, and infections that occurred since July 1, 2021. Residents and staff with a documented infection within the 90 days or a positive test within 30 days prior to the beginning of the study period were excluded because of difficulties discerning positive tests from breakthrough infections or past infections. Days since last infection is down in Table S3.

*Week*. Dates since the start of the study period were aggregated into 7-day intervals.

*Race or ethnic group*. We grouped race and ethnicity information, assigned by CDCR based on a combination of self-reports and administrative records, into categories used by the U.S. Census Bureau (Hispanic or Latine, non-Hispanic Black, non-Hispanic White, non-Hispanic Other or Unknown).^[3]^ Race or ethnic groupings were mutually exclusive.

*Covid-19 risk score*. The Covid-19 risk score was developed by CDCR to grade a resident’s risk of severe outcomes following SARS-CoV-2 infection. It is an index equal to the sum of weighted indicators for 17 items identified in the scientific literature as risk factors for severe Covid-19 outcomes.^[4]^ Residents who had Covid-19 risk scores of 2 or more were aged younger than 65 years with comorbidities or those aged 65 years and older (Table S1).

*Room type*. Residents are housed in rooms, discrete spaces that are at least partially enclosed by solid walls. We defined room type according to the number of residents housed in each room, dichotomizing this variable into cells (rooms with 1-2 occupants) and dormitories (rooms with 3 or more occupants).

*Age*. Age of individual in years.

*Gender*. Staff members self-reported their gender identity.

*Sex.* For residents, only sex was reported. We did not match for sex given strong collinearity with prison.

*Position*. We included staff members employed at a prison with a designation of custody or healthcare (excluding contract employees) and who worked in roles that involved regular direct contact with residents.

#### 2.5 Covid-19 outcomes and vaccination

The data included information about Covid-19 vaccination, testing, hospitalization, and death at the person-day record level.

*Vaccination.* Prison healthcare staff recorded each dose offer to residents, to whom it was made, and whether it was accepted or declined; this information was then entered into the CDCR electronic health record system. Residents who accepted a dose were vaccinated on the spot. Vaccination was available for all staff and was administered at each prison. Workers in specified correctional health care facilities had mandated vaccination requirements. Data for staff include vaccinations received in the community when logged in the California Immunization Registry and when the staff member gave permission for the registry data to be shared with other agencies. Cumulative vaccination of study cohort members, by vaccine status, is shown in Figure S3. Days since last vaccination is shown in Table S3.

*Confirmed infection.* A confirmed infection was defined as a positive real-time PCR or antigen diagnostic test for SARS-CoV-2. Positive tests were assigned to the date of specimen collection. Over the study period 99.9% of resident tests were by RT-PCR. All tests for staff were by RT-PCR.

*Hospitalization.* We defined hospitalization related to a SARS-CoV-2 infection as a hospitalization that occurred within three days prior to or 14 days after initial confirmation of an infection that included Covid-19 symptoms. Hospitalization and death data were only collected from residents. Counts within the study population for hospitalizations and deaths are shown in Table S3.

*Death*. All deaths related to a SARS-CoV-2 infection were classified and confirmed by the California Correctional Health Care Services.

### 3 Statistical analysis

#### 3.1 Matching methods

*Coarsened exact matching*

We used coarsened exact matching^[5]^ to match each case and control based on the case’s test week and vaccine type as well as relevant demographic, clinical, and location characteristics. Since individuals were repeatedly tested, their negative tests prior to censoring or study end could be matched as controls.

The following variables were used for the coarsened exact matching approach for each population:

Residents

- Week (categorical)
- Prison (categorical: 35 prisons)
- Covid-19 risk score (0, 1, ≥2)
- Room type (cell, dorm)

Staff

- Week (categorical)
- Prison (categorical: 35 prisons)
- Age (18-39, 40-54, ≥55)
- Position (custody, healthcare)
- Gender (male, female)

*Propensity score matching*

We conducted a sensitivity analysis in which we used propensity score matching as an alternative to coarsened exact matching. We included an expanded subset of demographic and medical characteristics that were previously identified as being predictive of vaccine acceptance among incarcerated people and staff.^[6],^^[7]^

Residents

- Week (categorical) [exact matching]
- Prison (categorical: 35 prisons) [exact matching]
- Covid-19 risk score (categorical: 0, 1, ≥2)
- Age (continuous)
- Room type (cell, dorm)
- Security level (categorical: 1, 2, 3, 4)
- Race or ethnic group (Hispanic or Latine, non-Hispanic Black, non-Hispanic white, non-Hispanic Other) [exact matching]

Staff

- Week (categorical) [exact matching]
- Prison (categorical: 35 prisons) [exact matching]
- Age (continuous)
- Position (custody, healthcare)
- Gender (male, female)
- Race or ethnic group (Hispanic or Latine, non-Hispanic Black, non-Hispanic white, non-Hispanic Other) [exact matching]

Nearest neighbor propensity score matching with replacement was used to match cases with controls in a 1:10 ratio. For variables marked as “exact matching,” propensity score matching was undertaken within strata defined by unique combinations of those variables.

#### 3.2 Regression models and estimands

The primary outcome of interest was SARS-CoV-2 infection, confirmed by positive RT-PCR or antigen test. Our main objective is to estimate the protection from variables related to vaccine- and infection-induced immunity. Variables were specified to capture protection from vaccination with at least two mRNA doses compared to no vaccination, and incremental protection from a third dose; similarly, we specified variables to capture protection from having at least one documented infection prior to the study start date, compared to no documented infection, and incremental protection from recency of infection, defined as the infection occurring since July 1, 2021. We included additional interacted terms for vaccine status and prior infection status. Our main rationale for including these terms were prior indications of interactions in neutralization assays.^[8]^

We estimated weighted conditional logistic regressions with the main predictors included as indicators and weights derived from coarsened exact matching. Odds ratios for the total estimands of vaccine and prior infection statuses were calculated as the product of odds ratios for individual and interacted covariates. Total estimands for immune status (unvaccinated & infected before July 1, 2021, unvaccinated & infected since July 1, 2021, vaccinated with two doses & no known infections, vaccinated with two doses & infected before July 1, 2021, vaccinated with two doses & infected since July 1, 2021, vaccinated with three doses & no known infections, vaccinated with three doses & infected before July 1, 2021, vaccinated with three doses & infected since July 1, 2021) were calculated as the sum of the coefficients for individual and interacted indictors and were defined relative to the unvaccinated & no known prior infections group.

Means and 95% confidence intervals were estimated using were calculated using 10,000 bias-corrected accelerated bootstrapped samples. Odds ratios of the total estimands of vaccine and prior infection statuses were calculated as the product of odds ratios for individual and interacted covariates. Total estimands of protection were calculated as 1 minus the odds ratio.

For the analyses that used propensity score matching, robust standard errors were used to calculate 95% confidence intervals given we match with replacement.^[9]^ The statistical model used on the matched propensity score used fully interacted terms between vaccine and prior infection status.

Analyses were performed using R software, version 3.5.2 (R Foundation for Statistical Computing).

#### 3.3 Sensitivity analyses

We conducted several sensitivity analyses (Tables S4 & S5) as follows:

A. Main study sample:
   i. Model adjusting for vaccine type
   ii. Stratified analysis by population (residents, staff)
B. Reducing the time-interval defining recent infections (shortening from 90 days to 30 days)
C. Excluding vaccinated persons who were not yet eligible for a third vaccine dose
D. Using inverse propensity score matching during the matching stage

### 4 Ethics approval

The study was approved by the institutional review board (IRB) at Stanford University (protocol #55835). The IRB approval of the study included a waiver of consent, on the basis that CDCR provided the Stanford research team with a limited data set without direct identifiers, the data had been collected for operational purposes, and the study could not practicably be carried out otherwise.

### 5 Supplementary tables and figures

#### Table S1. Covid-19 testing strategies and definitions for the resident testing program.^[10]^

| Strategy | Individuals Tested | Goals of Testing | Timing and Frequency |
| --- | --- | --- | --- |
| Diagnostic testing | • Patients with symptoms consistent with COVID-19 | • Confirm diagnosis of COVID-19 | • Test when patient presents with symptoms |
| Outbreak response testing | • Exposed inmates and staff who are susceptible to infection (naïve or >90 days since first positive test) • Close contacts of a confirmed case of COVID-19 (e.g., housing, yard, worksite, shared ventilation) • Broad-based testing of asymptomatic inmates in the entire facility or institution | • Determine the extent of transmission in the institution • Identify patients who are infected for isolation and monitoring • Limit transmission with rapid isolation | • As soon as possible after one or more cases are identified among inmates or staff at an institution • Serial testing every 3-7 days of groups (e.g., housing, worksite) of exposed inmates who initially test negative until all tests are negative in a 14-day time frame |
| Quarantine Testing | • Asymptomatic close contacts of a confirmed case of COVID-19 (e.g., housing, yard, worksite) in quarantine • Other asymptomatic exposed inmates in quarantine | • Identify patients who are infected for isolation and monitoring • Identify inmate contacts eligible for release from quarantine | • At the beginning of quarantine (within 24 hours if not tested within last 7 days or at 3 days post-exposure), middle of quarantine, and end of quarantine (day 12-14) trial testing every 3-7 days of all susceptible inmate contacts who initially test negative until all tests are negative in a 14-day time frame |
| Risk-based routine testing | • Patients using aerosol-generating devices (e.g., nebulizers or continuous positive airway pressure [CPAP] devices) • Vulnerable patients (age >65, comorbidities, COVID-19 weighted risk score ≥3) | • Early diagnosis to reduce morbidity and mortality among patients at higher risk of severe COVID-19 | • At least monthly in facilities or institutions in non-outbreak settings (no new cases in 14 days) • Upon patient request |
| Inmate-worker routine testing | • Susceptible inmates working in enclosed spaces with staff or inmates outside of their housing unit, in healthcare or other areas where patients are isolated or quarantined for COVID-19, providing health aid or peer support, or in jobs that require movement about the facility or institution | • Identify infections in asymptomatic non-exposed inmate workers to prevent transmission to other inmates and staff | • Weekly testing offer for susceptible inmate workers in non-outbreak settings |
| Testing for movement | • Arrivals from a county jail, return from out-to-court • Inter-facility transfers, transfers to fire camps • Release to the community, parole, probation • Inter-facility transfers for mental health, medical, or dental services • Returns from medical/hospital, off-site appointments | • Improve the safety of transfers • Reduce the risk of introduction of the virus • Early identification of asymptomatic infection | Procedures for testing during movement vary. Details can be found in California Correctional Health Care Services Covid-19 Interim Guidance: Covid-19 screening and testing matrix for patient movement |
| Public health surveillance testing | • Susceptible individuals representing cohorts of patients that regularly comingle or share air space (e.g., housing unit, worksite), excluding quarantined patients | • Detect outbreaks in an early phase to limit spread and prevent morbidity and mortality • Needed for the transition to higher re-opening phase | • Weekly testing of a sample of susceptible patients from each identified cohort in non-outbreak settings • Test 25% (up to 25) patients per cohort each week |

#### Table S2. Covid-19 risk score components and weights

| Condition | Definition | Weighted Score |
| --- | --- | --- |
| Age 65+ | Chronologic age of 65 years or above | 4 |
| Advanced liver disease | Advanced liver disease (cirrhosis/end stage liver disease) | 2 |
| Asthma | Persistent asthma (moderate or severe) as defined by the  California Correctional Health Care Services (CCHCS) asthma condition specifications | 1 |
| Cancer | High risk cancer as defined by the CCHCS cancer condition specifications (excludes most diagnoses of skin cancer and “personal history of” cancers”) | 2 |
| Chronic Kidney Disease | Chronic kidney disease as defined by the CCHCS chronic kidney disease condition specifications | 1 |
| Advanced Chronic Kidney Disease/Renal Failure | Chronic kidney disease (Stage 5) as defined by the CCHCS Chronic Kidney Disease Condition Specifications OR currently receiving Hemodialysis | 1 |
| Chronic Lung Disease (other) | Cystic fibrosis, pneumoconiosis, or pulmonary fibrosis | 1 |
| COPD | Chronic obstructive pulmonary disease | 2 |
| Diabetes | Diabetes | 1 |
| Diabetes (high risk) | High risk diabetes as defined by the CCHCS diabetes condition specifications | 1 |
| Heart Disease | Any of the following cardiovascular disease conditions: cerebrovascular, congestive heart failure, congenital heart disease, ischemic heart disease, peripheral vascular disease, thromboembolic disease, valvular disease, and cardiovascular disease (not otherwise specified). | 1 |
| Heart Disease (high risk) | High risk heart disease as defined by CCHCS condition specifications | 1 |
| Hemoglobin Disorder | Hemoglobin disorders as defined by CCHCS Condition Specifications for Hemoglobinopathy, including sickle cell disorder. | 1 |
| HIV | HIV | 1 |
| HIV (poorly controlled) | HIV with a CD4 count < 200 | 1 |
| Hypertension | Hypertension as defined by the CCHCS Hypertension Condition Specifications | 1 |
| Immunocompromised | Any of the following conditions: aplastic anemia, histiocytosis, immunosuppressed, organ transplant, other transplant | 2 |
| Neurologic conditions | Dementia, Parkinson's disease, multiple sclerosis, myasthenia gravis, or neurologic disorder as defined by CCHCS Condition Specifications | 1 |
| Obesity | Body mass index of 30 or above | 1 |
| Other high risk chronic condition | Any of the following conditions when they are high risk per CCHCS condition specifications: coccidioidomycosis, connective tissue disorder, endocrine disorder, or vasculitis | 1 |
| Pregnant | Pregnant | 1 |

#### Figure S1. Testing and cases among study cohort.

In the top panel, the daily number and percentage of residents tested and who were positive. In the bottom panel, the daily number of residents who tested positive. Testing and case series were extended over the historical period 2.5 months before the start of the study period. Staff were generally not tested during Federal holidays.

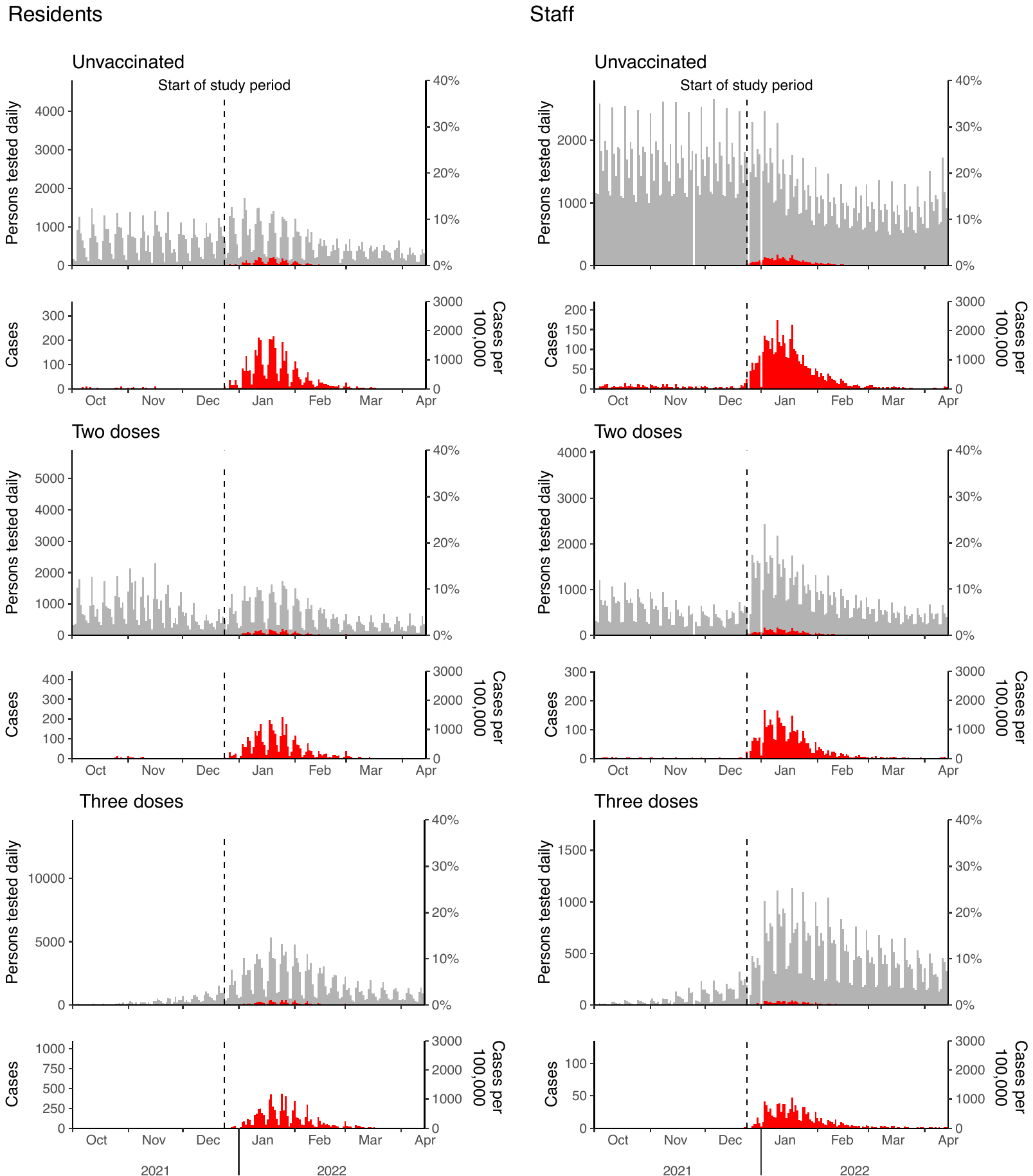

#### Table S3. Characteristics by vaccine and prior infection status.

Counts, testing, and timing of vaccination and prior infections. With the exception of counts for controls, summary statistics were not weighted. Controls are rounded to the nearest integer.

A. Main analysis against confirmed infection (90-day time interval for defining recent prior infections).

|  |  | **Residents** | | | | | | | | |
| --- | --- | --- | --- | --- | --- | --- | --- | --- | --- | --- |
| **Prior infection status** | **Vaccine status** | **Persons*** | **Cases** | **(%)^†^** | **Controls (weighted)** | **Hospitalizations** | **Deaths** | **Days since last infection (IQR)** | **Days since last vaccination (IQR)** | **Weekly test frequency^††^ (mean, sd)** |
| No known | Unvaccinated | 8766 | 3307 | 37.7% | 25429 | 46 | 0 | — | — | 0.8 (0.4) |
|  | Two doses | 9778 | 2810 | 28.7% | 25236 | 30 | 0 | — | 201 [104-316] | 0.7 (0.4) |
|  | Three doses | 19930 | 4482 | 22.5% | 51249 | 58 | 2 | — | 70 [51-95] | 0.8 (0.3) |
| Infected before July 1, 2021 | Unvaccinated | 2893 | 819 | 28.3% | 8287 | 4 | 0 | 415 [390-464] | — | 0.8 (0.4) |
|  | Two doses | 4826 | 1016 | 21.1% | 13983 | 8 | 0 | 417 [392-467] | 256 [132.75-297] | 0.7 (0.3) |
|  | Three doses | 17188 | 3246 | 18.9% | 53381 | 21 | 0 | 424 [397-483] | 64 [48-88] | 0.7 (0.3) |
| Infected since July 1, 2021 | Unvaccinated | 336 | 61 | 18.2% | 958 | 0 | 0 | 135 [110-167] | — | 0.8 (0.3) |
|  | Two doses | 223 | 27 | 12.1% | 796 | 0 | 0 | 150.5 [123-175] | 136.5 [66-286.25] | 0.7 (0.3) |
|  | Three doses | 299 | 15 | 5.0% | 848 | 1 | 0 | 158 [131-178] | 68 [50-85] | 0.8 (0.3) |
| Total |  | 64239 | 15783 | 24.6% | 180168 | 168 | 2 | 420 [392-475] | 77 [53-125] | 0.8 (0.4) |

|  |  | **Staff** | | | | | | |
| --- | --- | --- | --- | --- | --- | --- | --- | --- |
| **Prior infection status** | **Vaccine status** | **Persons*** | **Cases** | **(%)^†^** | **Controls (weighted)** | **Days since last infection (IQR)** | **Days since last vaccination (IQR)** | **Weekly test frequency^††^ (mean, sd)** |
| No known | Unvaccinated | 3781 | 2265 | 59.9% | 17879 | — | — | 1.8 (0.4) |
|  | Two doses | 7097 | 2785 | 39.2% | 24048 | — | 236 [117-336] | 1.4 (0.5) |
|  | Three doses | 4667 | 915 | 19.6% | 13994 | — | 61 [34-90] | 1.3 (0.5) |
| Infected before July 1, 2021 | Unvaccinated | 2281 | 1267 | 55.5% | 12357 | 405 [372-444] | — | 1.8 (0.4) |
|  | Two doses | 2436 | 715 | 29.4% | 9530 | 406 [378-453] | 157 [104-292] | 1.4 (0.5) |
|  | Three doses | 907 | 160 | 17.6% | 2831 | 416 [387-468] | 52 [28-78] | 1.3 (0.5) |
| Infected since July 1, 2021 | Unvaccinated | 1312 | 335 | 25.5% | 6861 | 153 [126-179] | — | 1.6 (0.5) |
|  | Two doses | 611 | 88 | 14.4% | 2467 | 159 [133-183] | 116 [76-269] | 1.4 (0.5) |
|  | Three doses | 176 | 9 | 5.1% | 440 | 160 [132-184] | 49 [27-76] | 1.2 (0.5) |
| Total |  | 23268 | 8539 | 36.7% | 90406 | 381 [192-427] | 114 [64-267] | 1.6 (0.5) |

B. Sensitivity analysis (30-day time interval for defining recent prior infections).

|  |  | **Residents** | | | | | | | | |
| --- | --- | --- | --- | --- | --- | --- | --- | --- | --- | --- |
| **Prior infection status** | **Vaccine status** | **Persons*** | **Cases** | **(%)^†^** | **Controls (weighted)** | **Hospitalizations** | **Deaths** | **Days since last infection (IQR)** | **Days since last vaccination (IQR)** | **Weekly test frequency^††^ (mean, sd)** |
| No known | Unvaccinated | 8766 | 3307 | 37.7% | 25344 | 46 | 0 | — | — | 0.8 (0.4) |
|  | Two doses | 9778 | 2810 | 28.7% | 25143 | 30 | 0 | — | 201 [104-316] | 0.7 (0.4) |
|  | Three doses | 19930 | 4482 | 22.5% | 51175 | 58 | 2 | — | 70 [51-95] | 0.8 (0.3) |
| Infected before July 1, 2021 | Unvaccinated | 2892 | 818 | 28.3% | 8276 | 4 | 0 | 415 [390-464] | — | 0.8 (0.4) |
|  | Two doses | 4826 | 1016 | 21.1% | 13977 | 8 | 0 | 417 [392-466] | 256 [132.75-297] | 0.7 (0.3) |
|  | Three doses | 17188 | 3246 | 18.9% | 53369 | 21 | 0 | 424 [397-483] | 64 [48-88] | 0.7 (0.3) |
| Infected since July 1, 2021 | Unvaccinated | 419 | 82 | 19.6% | 1383 | 0 | 0 | 117 [92-158] | — | 0.8 (0.3) |
|  | Two doses | 272 | 33 | 12.1% | 1000 | 0 | 0 | 139 [103-168] | 147 [69-309] | 0.7 (0.3) |
|  | Three doses | 346 | 16 | 4.6% | 1099 | 1 | 0 | 151 [104-175] | 63 [46-83] | 0.8 (0.4) |
| Total |  | 64417 | 15810 | 24.5% | 180766 | 168 | 2 | 419 [392-473] | 77 [53-125] | 0.8 (0.4) |

|  |  | **Staff** | | | | | | |
| --- | --- | --- | --- | --- | --- | --- | --- | --- |
| **Prior infection status** | **Vaccine status** | **Persons*** | **Cases** | **(%)^†^** | **Controls (weighted)** | **Days since last infection (IQR)** | **Days since last vaccination (IQR)** | **Weekly test frequency^††^ (mean, sd)** |
| No known | Unvaccinated | 3781 | 2265 | 59.9% | 17863 | — | — | 1.8 (0.4) |
|  | Two doses | 7097 | 2785 | 39.2% | 24039 | — | 236 [117-336] | 1.4 (0.5) |
|  | Three doses | 4668 | 915 | 19.6% | 14003 | — | 61 [34-90] | 1.3 (0.5) |
| Infected before July 1, 2021 | Unvaccinated | 2281 | 1267 | 55.5% | 12340 | 404 [372-444] | — | 1.8 (0.4) |
|  | Two doses | 2435 | 715 | 29.4% | 9523 | 406 [377.25-453] | 157 [104-292] | 1.4 (0.5) |
|  | Three doses | 906 | 160 | 17.7% | 2834 | 416 [386-468] | 52 [28-78] | 1.3 (0.5) |
| Infected since July 1, 2021 | Unvaccinated | 1340 | 354 | 26.4% | 7159 | 150 [122-178] | — | 1.6 (0.5) |
|  | Two doses | 646 | 94 | 14.6% | 2594 | 157 [128-181] | 119 [77-300] | 1.4 (0.5) |
|  | Three doses | 185 | 10 | 5.4% | 475 | 156 [124-182] | 49 [26.25-78] | 1.2 (0.5) |
| Total |  | 23339 | 8565 | 36.7% | 90830 | 380 [188-426] | 114 [64-268] | 1.6 (0.5) |

C. Sensitivity analysis (Exclusion of people vaccinated with two doses but were ineligible for a third dose).

|  |  | **Residents** | | | | | | | | |
| --- | --- | --- | --- | --- | --- | --- | --- | --- | --- | --- |
| **Prior infection status** | **Vaccine status** | **Persons*** | **Cases** | **(%)^†^** | **Controls (weighted)** | **Hospitalizations** | **Deaths** | **Days since last infection (IQR)** | **Days since last vaccination (IQR)** | **Weekly test frequency^††^ (mean, sd)** |
| No known | Unvaccinated | 8728 | 3307 | 37.9% | 26525 | 46 | 0 | — | — | 0.8 (0.4) |
|  | Two doses | 6145 | 1734 | 28.2% | 13782 | 19 | 0 | — | 307 [237-334] | 0.7 (0.4) |
|  | Three doses | 19878 | 4482 | 22.5% | 51710 | 58 | 2 | — | 70 [51-95] | 0.8 (0.3) |
| Infected before July 1, 2021 | Unvaccinated | 2889 | 819 | 28.3% | 8437 | 4 | 0 | 415 [390-464] | — | 0.8 (0.4) |
|  | Two doses | 3608 | 815 | 22.6% | 9699 | 6 | 0 | 418 [393-463] | 279 [247-309] | 0.7 (0.3) |
|  | Three doses | 17147 | 3246 | 18.9% | 53684 | 21 | 0 | 424 [397-483] | 64 [48-88] | 0.7 (0.3) |
| Infected since July 1, 2021 | Unvaccinated | 335 | 61 | 18.2% | 989 | 0 | 0 | 135 [110-166] | — | 0.8 (0.3) |
|  | Two doses | 104 | 11 | 10.6% | 387 | 0 | 0 | 151 [117.75-179.25] | 297 [188.75-340] | 0.7 (0.3) |
|  | Three doses | 298 | 15 | 5.0% | 873 | 1 | 0 | 158 [131-178] | 68 [50-85] | 0.8 (0.3) |
| Total |  | 59132 | 14490 | 24.5% | 166086 | 155 | 2 | 420 [393-475] | 77 [54-127] | 0.8 (0.3) |

|  |  | **Staff** | | | | | | |
| --- | --- | --- | --- | --- | --- | --- | --- | --- |
| **Prior infection status** | **Vaccine status** | **Persons*** | **Cases** | **(%)^†^** | **Controls (weighted)** | **Days since last infection (IQR)** | **Days since last vaccination (IQR)** | **Weekly test frequency^††^ (mean, sd)** |
| No known | Unvaccinated | 3776 | 2263 | 59.9% | 17848 | — | — | 1.8 (0.4) |
|  | Two doses | 5188 | 1816 | 35.0% | 14339 | — | 327 [250-349] | 1.4 (0.5) |
|  | Three doses | 4602 | 914 | 19.9% | 13532 | — | 61 [34-90] | 1.3 (0.5) |
| Infected before July 1, 2021 | Unvaccinated | 2281 | 1266 | 55.5% | 12324 | 404 [372-443] | — | 1.8 (0.4) |
|  | Two doses | 1621 | 465 | 28.7% | 4668 | 409 [380-462] | 280 [217-336] | 1.4 (0.5) |
|  | Three doses | 892 | 160 | 17.9% | 2764 | 416 [387-465] | 52 [28-78] | 1.3 (0.5) |
| Infected since July 1, 2021 | Unvaccinated | 1311 | 335 | 25.6% | 6853 | 152 [126-179] | — | 1.6 (0.5) |
|  | Two doses | 302 | 26 | 8.6% | 814 | 153 [123-185] | 331 [229.75-359] | 1.4 (0.5) |
|  | Three doses | 173 | 9 | 5.2% | 435 | 160 [131.75-184] | 49 [27-76] | 1.2 (0.5) |
| Total |  | 20146 | 7254 | 36.0% | 73577 | 381 [190-427] | 154 [60-322] | 1.6 (0.5) |

D. Sensitivity analysis (propensity score matching).

|  |  | **Residents** | | | | | | | | |
| --- | --- | --- | --- | --- | --- | --- | --- | --- | --- | --- |
| **Prior infection status** | **Vaccine status** | **Persons*** | **Cases** | **(%)^†^** | **Controls** | **Hospitalizations** | **Deaths** | **Days since last infection (IQR)** | **Days since last vaccination (IQR)** | **Weekly test frequency^††^ (mean, sd)** |
| No known | Unvaccinated | 7206 | 3309 | 45.9% | 22908 | 47 | 0 | — | — | 0.8 (0.3) |
|  | Two doses | 7515 | 2812 | 37.4% | 22586 | 30 | 0 | — | 158 [83-299] | 0.8 (0.3) |
|  | Three doses | 15497 | 4485 | 28.9% | 44099 | 58 | 2 | — | 63 [47-85] | 0.8 (0.3) |
| Infected before July 1, 2021 | Unvaccinated | 2354 | 819 | 34.8% | 7355 | 4 | 0 | 416 [392-474] | — | 0.8 (0.3) |
|  | Two doses | 3813 | 1017 | 26.7% | 12417 | 8 | 0 | 418 [393-478] | 256 [125-291] | 0.7 (0.3) |
|  | Three doses | 13823 | 3247 | 23.5% | 45980 | 21 | 0 | 430 [400-491] | 57 [45-77] | 0.7 (0.3) |
| Infected since July 1, 2021 | Unvaccinated | 234 | 61 | 26.1% | 938 | 0 | 0 | 122 [107-155] | — | 0.8 (0.3) |
|  | Two doses | 160 | 27 | 16.9% | 711 | 0 | 0 | 138 [113-166] | 113 [55-265] | 0.8 (0.3) |
|  | Three doses | 193 | 15 | 7.8% | 808 | 1 | 0 | 153 [127.5-173] | 55 [47-70] | 0.8 (0.3) |
| Total |  | 50795 | 15792 | 31.1% | 157802 | 169 | 2 | 423 [394-485] | 69 [48-116] | 0.8 (0.3) |

|  |  | **Staff** | | | | | | |
| --- | --- | --- | --- | --- | --- | --- | --- | --- |
| **Prior infection status** | **Vaccine status** | **Persons*** | **Cases** | **(%)^†^** | **Controls** | **Days since last infection (IQR)** | **Days since last vaccination (IQR)** | **Weekly test frequency^††^ (mean, sd)** |
| No known | Unvaccinated | 3746 | 2267 | 60.5% | 16561 | — | — | 1.8 (0.4) |
|  | Two doses | 6609 | 2790 | 42.2% | 22811 | — | 231 [109-335] | 1.4 (0.5) |
|  | Three doses | 4110 | 919 | 22.4% | 13083 | — | 55 [28-76] | 1.3 (0.5) |
| Infected before July 1, 2021 | Unvaccinated | 2271 | 1267 | 55.8% | 11652 | 398 [370-429] | — | 1.8 (0.4) |
|  | Two doses | 2271 | 717 | 31.6% | 9157 | 399 [374.25-439] | 156 [100-295] | 1.4 (0.5) |
|  | Three doses | 786 | 161 | 20.5% | 2669 | 407 [382-469] | 48 [25-69] | 1.3 (0.5) |
| Infected since July 1, 2021 | Unvaccinated | 1169 | 336 | 28.7% | 6724 | 146 [122-168] | — | 1.7 (0.4) |
|  | Two doses | 528 | 89 | 16.9% | 2320 | 154 [133-175] | 107 [66-262] | 1.4 (0.5) |
|  | Three doses | 136 | 9 | 6.6% | 422 | 149 [121-174] | 42 [21-65] | 1.2 (0.5) |
| Total |  | 21626 | 8555 | 39.6% | 85399 | 380 [189-416] | 111 [63-279] | 1.6 (0.5) |

* Total persons that contributed tests to study sample. Since prior infection and vaccine statuses were time-varying, counts were calculated relative to a persons’ last sample.

^†^ Percent positive was calculated relative to the total number of people in each strata.

^††^ Weekly test frequency was calculated as 7 × the number of tests over the study period divided by the number of days from the study start date to the last date of observation, defined as the first of the following: date of test collection for a positive test, date of release or death for residents, or last shift for staff.

#### Figure S2. Cumulative number of tests by person.

The number of cumulative tests as a proportion of people over the study period.

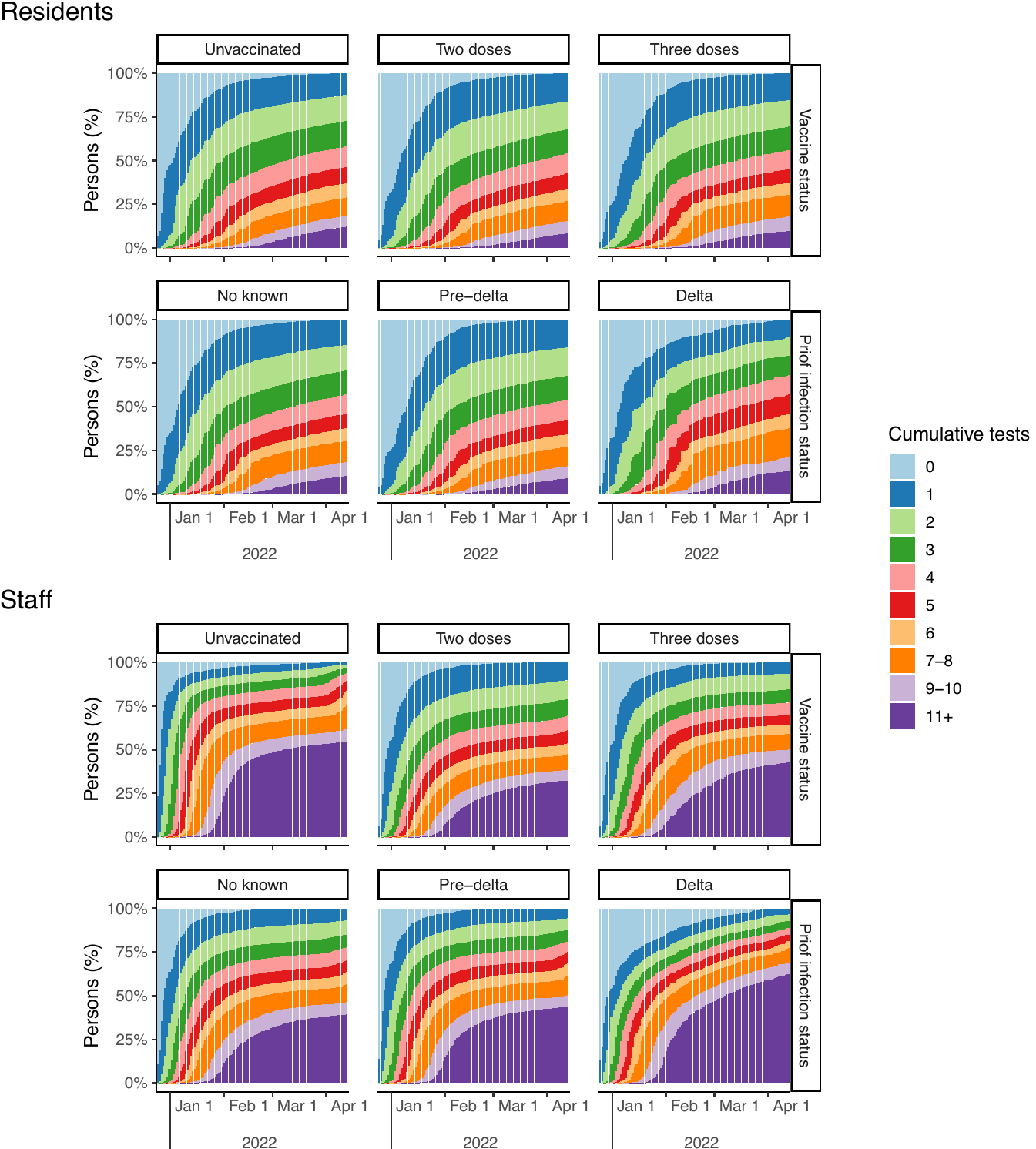

#### Figure S3. Distribution of prior infections among study cohort by vaccination status.

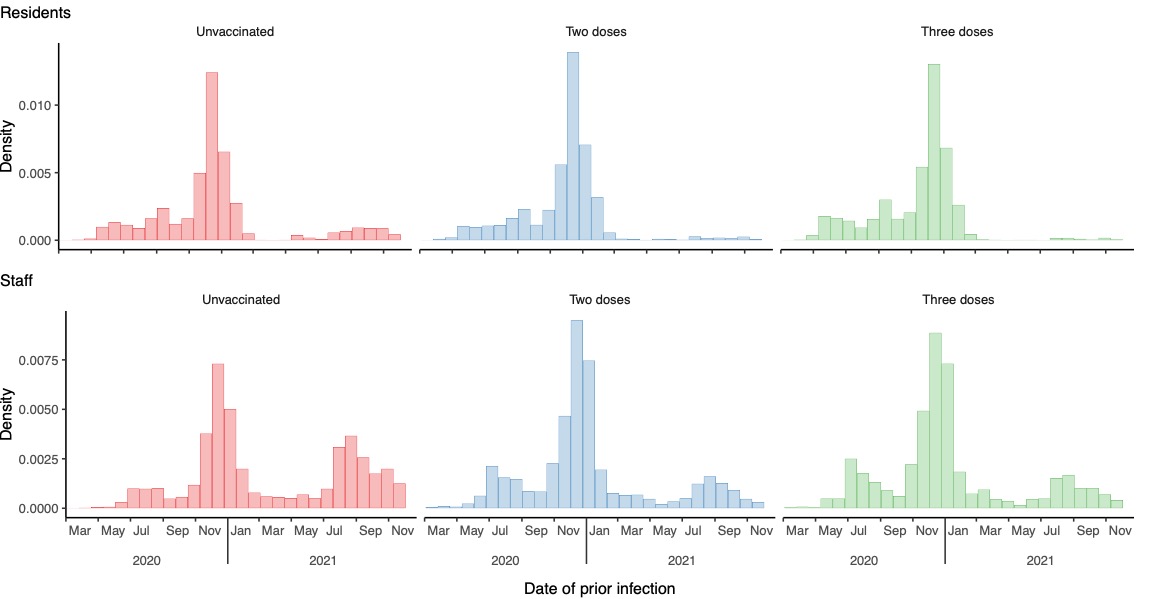

#### Table S4. Odds ratios from sensitivity analyses.

Odds ratios are calculated as the exponentiated coefficients derived from the logistic regression models.

A. Odds ratio estimates in pooled and stratified populations.

|  | **Pooled** | | **Stratified** | |
| --- | --- | --- | --- | --- |
| **Covariates** | **Main analysis** | **Model with indicator for vaccine type** | **Residents** | **Staff** |
| Any prior infection | 0.743 (0.684 to 0.766) | 0.744 (0.684 to 0.766) | 0.665 (0.599 to 0.714) | 0.814 (0.729 to 0.850) |
| Infected since July 1, 2021 | 0.528 (0.457 to 0.578) | 0.528 (0.457 to 0.578) | 0.689 (0.509 to 0.899) | 0.480 (0.404 to 0.529) |
| Received at least two doses | 0.851 (0.803 to 0.877) | 0.841 (0.799 to 0.877) | 0.815 (0.754 to 0.852) | 0.887 (0.819 to 0.933) |
| Received a third dose | 0.668 (0.629 to 0.688) | 0.670 (0.630 to 0.689) | 0.718 (0.677 to 0.756) | 0.523 (0.454 to 0.542) |
| Infected before July 1, 2021 & Vaccinated with two doses only | 0.826 (0.763 to 0.895) | 0.827 (0.763 to 0.895) | 0.887 (0.799 to 1.004) | 0.814 (0.710 to 0.896) |
| Infected before July 1, 2021 & Vaccinated with three doses | 0.916 (0.865 to 1.003) | 0.915 (0.864 to 1.003) | 0.974 (0.891 to 1.085) | 1.099 (0.875 to 1.321) |
| Infected since July 1, 2021 & Vaccinated with two doses only | 0.807 (0.630 to 0.990) | 0.801 (0.628 to 0.988) | 0.672 (0.355 to 1.031) | 0.801 (0.606 to 1.018) |
| Infected since July 1, 2021 & Vaccinated with three doses | 0.594 (0.355 to 0.877) | 0.594 (0.354 to 0.876) | 0.468 (0.221 to 0.767) | 0.770 (0.271 to 1.414) |
| Pfizer |  | 1.044 (0.960 to 1.051) |  |  |

B. Odds ratio estimates in the pooled and stratified populations with the time-interval for recent infections shortened from 90 days to 30 days.

| **Covariates** | **Pooled** | **Residents** | **Staff** |
| --- | --- | --- | --- |
| Any prior infection | 0.744 (0.955 to 0.955) | 0.665 (0.600 to 0.715) | 0.815 (0.725 to 0.849) |
| Infected since July 1, 2021 | 0.534 (0.790 to 0.790) | 0.665 (0.501 to 0.829) | 0.487 (0.415 to 0.534) |
| Received at least two doses | 0.851 (0.802 to 0.875) | 0.815 (0.760 to 0.857) | 0.886 (0.818 to 0.930) |
| Received a third dose | 0.668 (0.594 to 0.623) | 0.718 (0.675 to 0.754) | 0.523 (0.451 to 0.540) |
| Infected before July 1, 2021 & Vaccinated with two doses only | 0.826 (0.785 to 0.972) | 0.887 (0.796 to 1.008) | 0.814 (0.711 to 0.903) |
| Infected before July 1, 2021 & Vaccinated with three doses | 0.915 (0.764 to 1.117) | 0.974 (0.890 to 1.091) | 1.099 (0.872 to 1.315) |
| Infected since July 1, 2021 & Vaccinated with two doses only | 0.800 (1.127 to 1.127) | 0.681 (0.395 to 1.010) | 0.803 (0.610 to 1.008) |
| Infected since July 1, 2021 & Vaccinated with three doses | 0.526 (1.416 to 1.416) | 0.405 (0.203 to 0.665) | 0.784 (0.314 to 1.417) |

C. Odds ratio estimates in the pooled and stratified populations for analysis excluding people vaccinated with two doses but were ineligible for a third dose.

| **Covariates** | **Pooled** | **Residents** | **Staff** |
| --- | --- | --- | --- |
| Any prior infection | 0.750 (0.689 to 0.775) | 0.675 (0.606 to 0.726) | 0.815 (0.730 to 0.854) |
| Infected since July 1, 2021 | 0.524 (0.457 to 0.579) | 0.678 (0.500 to 0.893) | 0.477 (0.406 to 0.530) |
| Received at least two doses | 0.949 (0.910 to 1.007) | 0.927 (0.871 to 1.001) | 0.961 (0.899 to 1.040) |
| Received a third dose | 0.610 (0.562 to 0.621) | 0.645 (0.593 to 0.674) | 0.489 (0.412 to 0.496) |
| Infected before July 1, 2021 & Vaccinated with two doses only | 0.886 (0.806 to 0.969) | 0.904 (0.796 to 1.028) | 0.985 (0.857 to 1.126) |
| Infected before July 1, 2021 & Vaccinated with three doses | 0.905 (0.852 to 0.990) | 0.959 (0.871 to 1.070) | 1.092 (0.880 to 1.338) |
| Infected since July 1, 2021 & Vaccinated with two doses only | 0.628 (0.364 to 0.822) | 0.503 (0.130 to 0.884) | 0.651 (0.359 to 0.901) |
| Infected since July 1, 2021 & Vaccinated with three doses | 0.580 (0.347 to 0.860) | 0.458 (0.215 to 0.748) | 0.758 (0.280 to 1.434) |

D. Odds ratio estimates in the pooled and stratified populations using propensity score matching.

| **Prior infection status** | **Vaccine status** | **Pooled** | **Residents** | **Staff** |
| --- | --- | --- | --- | --- |
| No known | Unvaccinated | *Ref.* | *Ref.* | *Ref.* |
|  | Two doses | 0.853 (0.820 to 0.888) | 0.810 (0.767 to 0.856) | 0.872 (0.820 to 0.926) |
|  | Three doses | 0.606 (0.580 to 0.632) | 0.592 (0.561 to 0.624) | 0.462 (0.423 to 0.504) |
| Infected before July 1, 2021 | Unvaccinated | 0.741 (0.702 to 0.782) | 0.659 (0.606 to 0.717) | 0.794 (0.738 to 0.855) |
|  | Two doses | 0.528 (0.498 to 0.559) | 0.473 (0.438 to 0.511) | 0.557 (0.509 to 0.609) |
|  | Three doses | 0.424 (0.404 to 0.446) | 0.382 (0.360 to 0.404) | 0.405 (0.342 to 0.479) |
| Infected since July 1, 2021 | Unvaccinated | 0.356 (0.320 to 0.396) | 0.413 (0.318 to 0.538) | 0.364 (0.323 to 0.409) |
|  | Two doses | 0.260 (0.216 to 0.314) | 0.241 (0.164 to 0.355) | 0.270 (0.217 to 0.336) |
|  | Three doses | 0.126 (0.084 to 0.189) | 0.108 (0.065 to 0.181) | 0.139 (0.072 to 0.270) |

#### Table S5. Total estimands from sensitivity analyses.

Estimate of protection for the multivariate logistic regression models. The joint protection for each prior infection and vaccine status combination was calculated as 1 minus the product of odds ratios for its corresponding covariates.

A. Estimated total protection in pooled and stratified populations.

|  |  | **Pooled** | | **Stratified** | |
| --- | --- | --- | --- | --- | --- |
| **Prior infection status** | **Vaccine status** | **Main analysis** | **Model with indicator for vaccine type** | **Residents** | **Staff** |
| No known | Unvaccinated | *Ref.* | *Ref.* | *Ref.* | *Ref.* |
|  | Two doses | 14.9% (12.3 to 19.7) | 15.9% (12.3 to 20.1) | 18.5% (14.8 to 24.6) | 11.3% (6.7 to 18.1) |
|  | Three doses | 43.2% (42.2 to 47.4) | 43.6% (42.2 to 47.5) | 41.5% (39.3 to 45.5) | 53.6% (52.3 to 60.5) |
| Infected before July 1, 2021 | Unvaccinated | 25.7% (23.4 to 31.6) | 25.6% (23.4 to 31.6) | 33.5% (28.6 to 40.1) | 18.6% (15.0 to 27.1) |
|  | Two doses | 47.8% (46.6 to 52.8) | 48.3% (46.6 to 52.9) | 52.0% (49.0 to 56.6) | 41.2% (39.7 to 50.1) |
|  | Three doses | 61.3% (60.7 to 64.8) | 61.7% (60.8 to 64.9) | 62.1% (60.8 to 65.2) | 58.4% (55.9 to 69.8) |
| Infected since July 1, 2021 | Unvaccinated | 60.8% (58.4 to 66.8) | 60.7% (58.4 to 66.8) | 54.2% (41.4 to 66.2) | 60.9% (58.5 to 67.8) |
|  | Two doses | 73.1% (69.8 to 80.1) | 73.6% (69.9 to 80.1) | 74.9% (66.1 to 86.7) | 72.2% (68.0 to 80.0) |
|  | Three doses | 86.8% (82.1 to 92.7) | 86.8% (82.1 to 92.7) | 87.4% (81.2 to 94.0) | 86.0% (77.2 to 95.9) |

B. Estimated total protection in the pooled and stratified populations with the time-interval for recent infections shortened from 90 days to 30 days.

| **Prior infection status** | **Vaccine status** | **Pooled** | **Residents** | **Staff** |
| --- | --- | --- | --- | --- |
| No known | Unvaccinated | *Ref.* | *Ref.* | *Ref.* |
|  | Two doses | 14.9% (12.5 to 19.8) | 15.9% (12.0 to 19.8) | 18.5% (14.3 to 24.0) |
|  | Three doses | 43.1% (47.7 to 50.1) | 43.6% (47.6 to 50.1) | 41.4% (39.2 to 45.5) |
| Infected before July 1, 2021 | Unvaccinated | 25.6% (4.5 to 4.5) | 25.6% (4.5 to 4.5) | 33.5% (28.5 to 40.0) |
|  | Two doses | 47.7% (29.8 to 30.3) | 48.3% (30.2 to 30.8) | 51.9% (48.8 to 56.5) |
|  | Three doses | 61.3% (42.5 to 59.0) | 61.6% (41.6 to 58.6) | 62.1% (60.6 to 65.0) |
| Infected since July 1, 2021 | Unvaccinated | 60.3% (39.1 to 39.1) | 60.3% (39.1 to 39.1) | 55.7% (46.1 to 66.9) |
|  | Two doses | 73.0% (51.8 to 51.8) | 73.4% (51.7 to 51.7) | 75.4% (67.7 to 85.7) |
|  | Three doses | 88.1% (61.2 to 61.2) | 88.2% (61.2 to 61.2) | 89.5% (84.7 to 94.9) |

C. Estimated total protection in the pooled and stratified populations for analysis excluding people vaccinated with two doses but were ineligible for a third dose.

| **Prior infection status** | **Vaccine status** | **Pooled** | **Residents** | **Staff** |
| --- | --- | --- | --- | --- |
| No known | Unvaccinated | *Ref.* | *Ref.* | *Ref.* |
|  | Two doses | 5.1% (-0.7 to 9.0) | 7.3% (-0.1 to 12.9) | 3.9% (-4.0 to 10.1) |
|  | Three doses | 42.1% (40.8 to 46.1) | 40.2% (37.5 to 44.3) | 53.0% (51.9 to 60.7) |
| Infected before July 1, 2021 | Unvaccinated | 25.0% (22.5 to 31.1) | 32.5% (27.4 to 39.4) | 18.5% (14.6 to 27.0) |
|  | Two doses | 36.9% (33.6 to 42.3) | 43.5% (38.8 to 48.7) | 22.9% (15.9 to 32.9) |
|  | Three doses | 60.7% (59.9 to 64.1) | 61.3% (59.7 to 64.5) | 58.2% (54.7 to 69.5) |
| Infected since July 1, 2021 | Unvaccinated | 60.7% (57.8 to 66.4) | 54.3% (41.1 to 66.6) | 61.1% (58.4 to 67.7) |
|  | Two doses | 76.6% (70.8 to 87.2) | 78.7% (64.7 to 94.2) | 75.7% (68.3 to 87.4) |
|  | Three doses | 86.8% (81.7 to 92.6) | 87.5% (81.5 to 94.0) | 86.1% (76.9 to 95.9) |

D. Estimated total protection in the pooled and stratified populations using propensity score matching.

| **Prior infection status** | **Vaccine status** | **Pooled** | **Residents** | **Staff** |
| --- | --- | --- | --- | --- |
| No known | Unvaccinated | *Ref.* | *Ref.* | *Ref.* |
|  | Two doses | 14.7% (11.2 to 18.0) | 19.0% (14.4 to 23.3) | 12.8% (7.4 to 18.0) |
|  | Three doses | 39.4% (36.8 to 42.0) | 40.8% (37.6 to 43.9) | 53.8% (49.6 to 57.7) |
| Infected before July 1, 2021 | Unvaccinated | 25.9% (21.8 to 29.8) | 34.1% (28.3 to 39.4) | 20.6% (14.5 to 26.2) |
|  | Two doses | 47.2% (44.1 to 50.2) | 52.7% (48.9 to 56.2) | 44.3% (39.1 to 49.1) |
|  | Three doses | 57.6% (55.4 to 59.6) | 61.8% (59.6 to 64.0) | 59.5% (52.1 to 65.8) |
| Infected since July 1, 2021 | Unvaccinated | 64.4% (60.4 to 68.0) | 58.7% (46.2 to 68.2) | 63.6% (59.1 to 67.7) |
|  | Two doses | 74.0% (68.6 to 78.4) | 75.9% (64.5 to 83.6) | 73.0% (66.4 to 78.3) |
|  | Three doses | 87.4% (81.1 to 91.6) | 89.2% (81.9 to 93.5) | 86.1% (73.0 to 92.8) |

### 6 STROBE Statement

#### 6.1 STROBE checklist of items that should be included in reports of cohort studies

|  | Item No | Recommendation | Page No^†^ |
| --- | --- | --- | --- |
| **Title and abstract** | 1 | (*a*) Indicate the study’s design with a commonly used term in the title or the abstract | p.2 |
|  |  | (*b*) Provide in the abstract an informative and balanced summary of what was done and what was found | p.2 |
| Introduction | | | |
| Background/rationale | 2 | Explain the scientific background and rationale for the investigation being reported | p.3 |
| Objectives | 3 | State specific objectives, including any prespecified hypotheses | p.3 |
| Methods | | | |
| Study design | 4 | Present key elements of study design early in the paper | p.4 |
| Setting | 5 | Describe the setting, locations, and relevant dates, including periods of recruitment, exposure, follow-up, and data collection | pp.4-5 |
| Participants | 6 | (*a*) Give the eligibility criteria, and the sources and methods of selection of participants. Describe methods of follow-up | pp.4-5, & Supp. Append. s.II |
|  |  | (*b*) For matched studies, give matching criteria and number of exposed and unexposed | pp.5-6 |
| Variables | 7 | Clearly define all outcomes, exposures, predictors, potential confounders, and effect modifiers. Give diagnostic criteria, if applicable | pp.4-5 & Supp. Append ss.II, III |
| Data sources/ measurement | 8^*^ | For each variable of interest, give sources of data and details of methods of assessment (measurement). Describe comparability of assessment methods if there is more than one group | p.4-5 & Supp. Append s.II |
| Bias | 9 | Describe any efforts to address potential sources of bias | pp.5-6 & Supp. Append. s.II, III |
| Study size | 10 | Explain how the study size was arrived at | p.4-5 |
| Quantitative variables | 11 | Explain how quantitative variables were handled in the analyses. If applicable, describe which groupings were chosen and why | pp.4-6 & Supp. Append. s.II, III |
| Statistical methods | 12 | (*a*) Describe all statistical methods, including those used to control for confounding | pp.5-6 & Supp. Append. s.III |
|  |  | (*b*) Describe any methods used to examine subgroups and interactions | pp.6 & Supp. Append. s.III |
|  |  | (*c*) Explain how missing data were addressed | p.4 & Supp. Append. s.II |
|  |  | (*d*) If applicable, explain how loss to follow-up was addressed | NA |
|  |  | (*e*) Describe any sensitivity analyses | p.6 |
| Results | | |  |
| Participants | 13^*^ | (a) Report numbers of individuals at each stage of study—eg numbers potentially eligible, examined for eligibility, confirmed eligible, included in the study, completing follow-up, and analysed | p.7, Figure 1 & Supp. Append. s.II |
|  |  | (b) Give reasons for non-participation at each stage | NA |
|  |  | (c) Consider use of a flow diagram | Figure 1 |
| Descriptive data | 14^*^ | (a) Give characteristics of study participants (eg demographic, clinical, social) and information on exposures and potential confounders | Table 1 |
|  |  | (b) Indicate number of participants with missing data for each variable of interest | Fig.1 |
|  |  | (c) Summarise follow-up time (eg, average and total amount) | p.7 & Table S3 |
| Outcome data | 15^*^ | Report numbers of outcome events or summary measures over time | p.7, Figures 1 & 2 |
| Main results | 16 | (*a*) Give unadjusted estimates and, if applicable, confounder-adjusted estimates and their precision (eg, 95% confidence interval). Make clear which confounders were adjusted for and why they were included | p.8, Figure 4 & Supp. Append. s.III |
|  |  | (*b*) Report category boundaries when continuous variables were categorized | p.8 |
|  |  | (*c*) If relevant, consider translating estimates of relative risk into absolute risk for a meaningful time period | NA |
| Other analyses | 17 | Report other analyses done—eg analyses of subgroups and interactions, and sensitivity analyses | p.8, Tables S4 and S5, & Supp. Append. s.III |
| Discussion | | | |
| Key results | 18 | Summarise key results with reference to study objectives | p.9 |
| Limitations | 19 | Discuss limitations of the study, taking into account sources of potential bias or imprecision. Discuss both direction and magnitude of any potential bias | pp.10-11 |
| Interpretation | 20 | Give a cautious overall interpretation of results considering objectives, limitations, multiplicity of analyses, results from similar studies, and other relevant evidence | pp.9-11 |
| Generalisability | 21 | Discuss the generalisability (external validity) of the study results | p.11 |
| Other information | | | |
| Funding | 22 | Give the source of funding and the role of the funders for the present study and, if applicable, for the original study on which the present article is based | p.11 |

^*^ Give information separately for exposed and unexposed groups.

^†^ Page numbers reflect the numbering of the final submitted version of manuscript.

1. Chin ET, Ryckman T, Prince L, Leidner D, Alarid-Escudero F, Andrews JR, Salomon JA, Studdert DM, Goldhaber-Fiebert JD. COVID-19 in the California State Prison System: an Observational Study of Decarceration, Ongoing Risks, and Risk Factors. J Gen Intern Med. 2021 Oct;36(10):3096-3102.
2. U.S. Centers for Disease Control and Prevention. COVID-19 Vaccination Clinical & Professional Resources. https://www.cdc.gov/vaccines/covid-19/index.html. Accessed: December 1, 2021.
3. U.S. Census Bureau. 2010 Census Summary File 1— Technical Documentation. Washington, DC: U.S. Department of Commerce. SF1/10-4 (RV).
4. Chin ET, Leidner D, Zhang Y, et al. Effectiveness of the mRNA-1273 Vaccine during a SARS-CoV-2 Delta Outbreak in a Prison. N Engl J Med. 2021;385(24):2300-2301. doi:10.1056/NEJMc2114089
5. Iacus SM, King G, Porro G. Causal inference without balance checking: coarsened exact matching. Polit anal. 2012;20(1):1–24.
6. Chin ET, Leidner D, Ryckman T, et al. Covid-19 Vaccine Acceptance in California State Prisons. N Engl J Med. 2021;385(4):374-376. doi:10.1056/NEJMc2105282.
7. Prince L, Long E, Studdert DM, et al. Uptake of COVID-19 Vaccination Among Frontline Workers in California State Prisons. JAMA Health Forum. 2022;3(3):e220099. doi:10.1001/jamahealthforum.2022.0099.
8. Gruell H, Vanshylla K, Tober-Lau P, et al. mRNA booster immunization elicits potent neutralizing serum activity against the SARS-CoV-2 Omicron variant [published online ahead of print, 2022 Jan 19]. Nat Med. 2022;1-4. doi:10.1038/s41591-021-01676-0.
9. Abadie A, Imbens GW. On the failure of the boostrap for matching estimators. Econometrica; 76(6). 2008. doi:10.3982/ECTA6474
10. California Correctional Health Care Services. COVID-19 Testing Definitions and Strategies. COVID-19 and Seasonal Influenza: Interim Guidance for Health Care and Public Health Providers. Revised: February 19, 2021.
